# IMPROVING HAND HYGIENE IN CRUISE SHIP: AN INTERVENTION STUDY

**DOI:** 10.1101/2025.03.22.25324349

**Authors:** Szava Bansaghi, EU HEALTHY SAILING project, Jörn Klein

## Abstract

Cruise ships are relatively small, crowded spaces where many people travel in a new environment, making infection control especially important. Proper hand hygiene is the first line of defense against the spread of infection. This study aimed to measure the effectiveness of different hand hygiene improvement measures onboard.

The study took place on the *Celestyal Olympia*, a medium-sized cruise ship. Our intervention study had four arms: first, baseline parameters were measured. Then, three different interventions were implemented—surface disinfection with antimicrobial spray, behavioral change through hand hygiene monitoring, and training.

Each person onboard used on average 7.6 soap doses and 1.6 hand-rub doses per day, indicating suboptimal hand hygiene frequency. Surface disinfection sprays were not proven to effectively reduce microbial loads on surfaces. Hand hygiene monitoring devices can only be effective if crew members can use them during their shifts. Designing hand hygiene training for crew members is challenging, as they have very limited preexisting knowledge of health sciences.

There were some additional lessons from the study. Hand hygiene compliance is primarily determined by the setting—the placement of dispensers and whether passengers are reminded to use them. Language barriers are a limiting factor that should be considered when planning communication strategies for both crew and passengers.

On cruise ships, hand hygiene is often associated with food hygiene. While there are clear recommendations in that area, there is a lack of guidelines on how to improve passengers’ hand hygiene. Some passengers have strong opinions about hand hygiene—either positive or negative—but they are a minority. The majority of passengers are not interested on hand hygiene; they perform it when the setting is optimal but otherwise skip it. Clear recommendations are needed to establish an environment that effectively promotes hand hygiene.

## INTRODUCTION

Cruise tourism has become increasingly popular. Outbreaks of infectious diseases are frequently reported aboard cruise ships [1]. Traveling on cruise ships exposes individuals to new environments and large numbers of people, increasing the risk of infection transmission [2]. Frequent personal interactions, complex population flows, limited space, and defective infrastructure aboard many cruise ships make them potential incubators for infectious diseases. Infection control on cruise ships is particularly challenging due to shared living and dining spaces, rapid turnover of passengers, and numerous opportunities for germs to be introduced on board [3].

Proper hand washing is the first line of defense against the spread of many illnesses on board [4]. In an outbreak investigation, more than 90% of passengers reported increasing their hand hygiene practices after becoming aware of the outbreak [3]. However, evidence-based strategies to proactively improve hand hygiene among cruise ship passengers (outside of outbreak scenarios) are not well established. Few studies have systematically tested interventions in this unique setting, leaving a gap in guidance for cruise operators.

This study was designed to assess the effectiveness of various interventions aimed at increasing hand hygiene practices using multiple objective, evidence-based evaluation methods. We hypothesized that implementing (1) antimicrobial surface treatments, (2) a technology-based hand hygiene feedback device, and (3) targeted crew training (each added to baseline conditions) would incrementally improve hand hygiene outcomes on the ship.

## METHODS

The intervention study aimed to investigate the impact of various infection-control measures on several outcomes (Table 1).

**Table 1:**
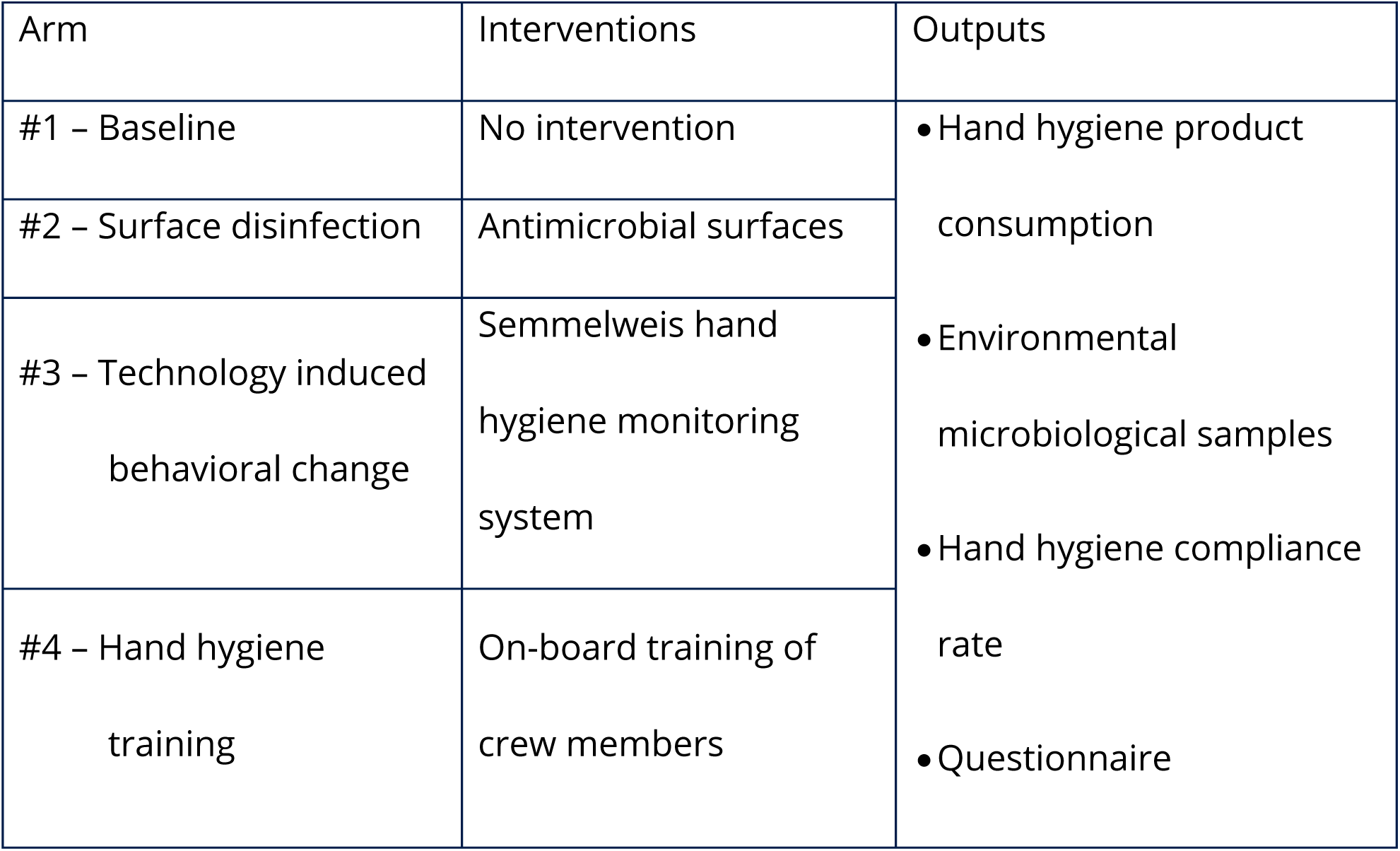
Structure of the intervention study.

### Interventions

The study had four arms. In the first arm, baseline data were collected. Each subsequent arm introduced a new element of infection control measures.

### Surface disinfection

Frequently touched surfaces are significant in infection transmission. Antimicrobial sprays are marketed as treatments that can make surfaces continuously self-disinfecting. That would be huge step forward in infection control, as these self-cleaning surfaces would break the infection transmission chain.

Two commercially available antimicrobial surface treatment sprays were selected. To protect the manufacturers’ commercial interests, the product names will not be disclosed. The active ingredients of these sprays were two different quaternary ammonium compounds; dimethyl-octadecyl-(3-trimethoxysilylpropyl)-azanium-chloride (DMOAP) in Spray#A, and a combination of didecyldimethylammonium chloride (DDAC) and benzalkonium chloride (BAC) in Spray#B. Spray #A claims that it reduces colony-forming units (CFU) by 90% compared to conventional cleaning, even after 60 days. Spray #B claims to kill 99.9999% of germs, and to keep surfaces bacteria-free for up to 30 days.

The two sprays were applied on several frequently touched surfaces of the ship: at the two main staircases, all inner rail, restroom knobs in Deck 4, 5 and 7. All outside and inside elevator panels were treated in case of the 6 main elevators.

### Technology-induced behavioral change

The Semmelweis Hand Hygiene System (HandInScan Zrt.) was installed on the ship as an innovative solution designed to visualize hand coverage after a hand hygiene. The system operates using the fluorescence-based method, one of the most widely applied techniques for assessing hand hygiene effectiveness. The handrub contains a fluorescent marker. After hand disinfection, under UV-A light, properly treated areas glow, while untreated areas remain dark. The Semmelweis System detects these covered (and theoretically disinfected) areas and provides immediate, objective feedback on hand hygiene performance (Figure 1).

Figure 1: The Semmelweis System installed in the buffet onboard, displaying feedback on properly disinfected hand surface areas.

*The original version of this figure included photographic materials that have been removed to comply with preprint policies prohibiting identifiable images. Readers interested in accessing the original visual data may contact the corresponding author for further information*.

Crew members were given QR codes to allow the system to identify them. The QR codes were assigned randomly. Participation was voluntary for all crew members; only a portion of the crew took part in the study. Passengers were also offered the opportunity to use the device but without identification.

### Hand Hygiene Training

Hand hygiene training sessions were conducted by USN researchers on October 30th, 31st, and November 2nd. Crew members attended in small groups of approximately 15 people each. The training sessions lasted between 20 and 25 minutes. Over the course of the three days, a total of 367 crew members participated. Crew were asked to complete a questionnaire assessing knowledge and attitudes, some before and some after the training, to gauge knowledge improvement (see Outcomes).

After the training, the latest version of the training material was recorded as a video for future use. The video was edited using Microsoft PowerPoint. It is available in other languages besides English. Translations were provided by ChatGPT, and the text was read aloud using NaturalReaders (https://www.naturalreaders.com/online/).

Meanwhile, passengers received a flyer emphasizing the importance of hand hygiene onboard (Figure 2). The flyer was distributed as a pillow letter, placed in passenger rooms upon arrival. It highlighted cruise-specific moments when hand hygiene is required and invited passengers to use the Semmelweis System to check their hand hygiene performance at any time.

**Figure 2:**
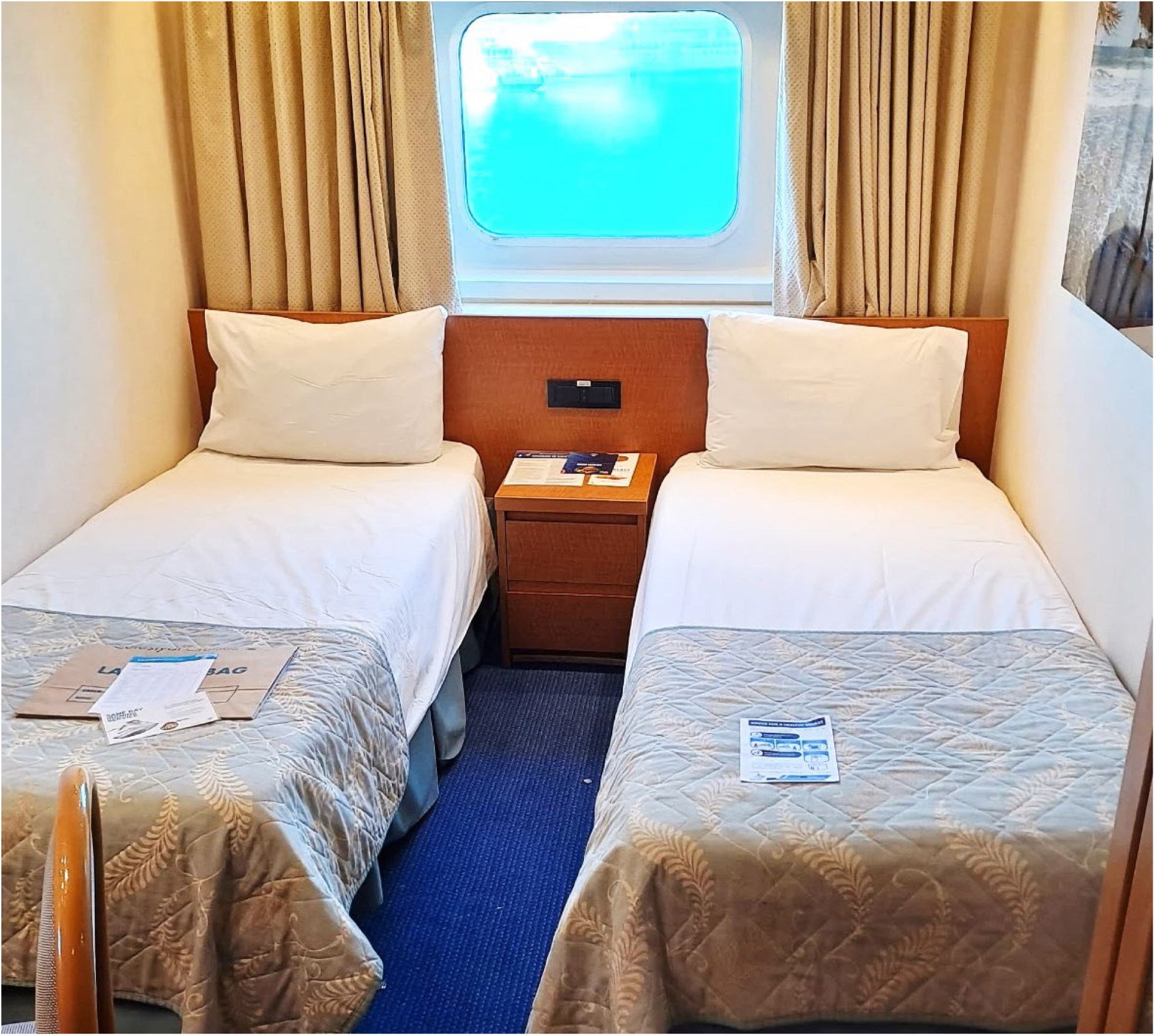
Flyer about hand hygiene for passengers, distributed as a pillow letter.

### Outcomes

#### Hand hygiene product consumption

Hand hygiene product consumption was calculated using the following data: usage records from the Provision Master’s database were compared to the number of guest nights provided by the F&B Manager’s database. The number of crew members remained relatively stable throughout the season, with data sourced from the Hotel Manager. The volume of product dispensed by onboard dispensers was also measured using the same method previously described [5].

#### Environmental microbiological samples

Eight frequently touched surfaces were sampled onboard, once during each Arm (Figure 3, Supplementary Table 1). Depending on the size and shape of the surfaces, samples were collected using tryptic soy agar (TSA) contact plates (VWR C31114TI), contact slides containing nutrient agar (NA) (VWR 535092Q), or FLOQ swabs (Copan) moistened with sterile 0.9% NaCl solution (R-Biopharm Z0301), and cultured on TSA plates (VWR 17114ZA). Samples were incubated at 35–36°C for 48 hours. After incubation, colony-forming units (CFUs) were counted. In each case, a maximum of 200 colonies were counted; any sample exceeding this number was labeled as “200+”. Traffic at the selected surfaces was also recorded, by direct observations.

**Figure 3:**
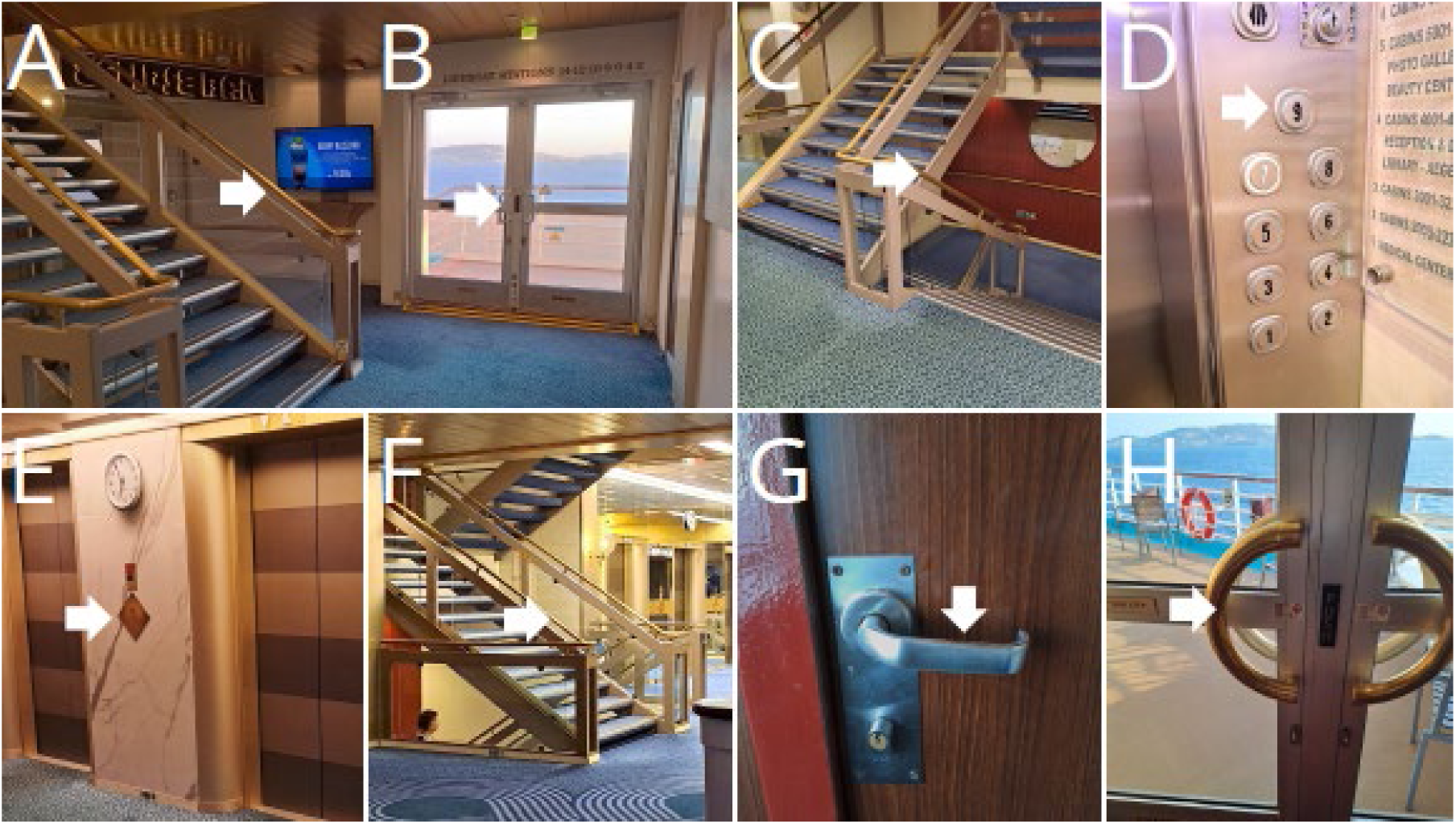
Selected surfaces for environmental sample collection.

#### Hand hygiene compliance rate

Hand hygiene compliance is a rate that tells us from all the opportunities when hand hygiene would be required how many times people actually performed it. Three key moments where selected where handwashing would be necessary and observation was feasible: entering the ship, leaving the ship, and entering the restaurants (Figure 4). There are additional critical moments for hand hygiene, (after using the restroom or upon entering a cabin), but these could not be observed due to privacy concerns.

**Figure 4:**
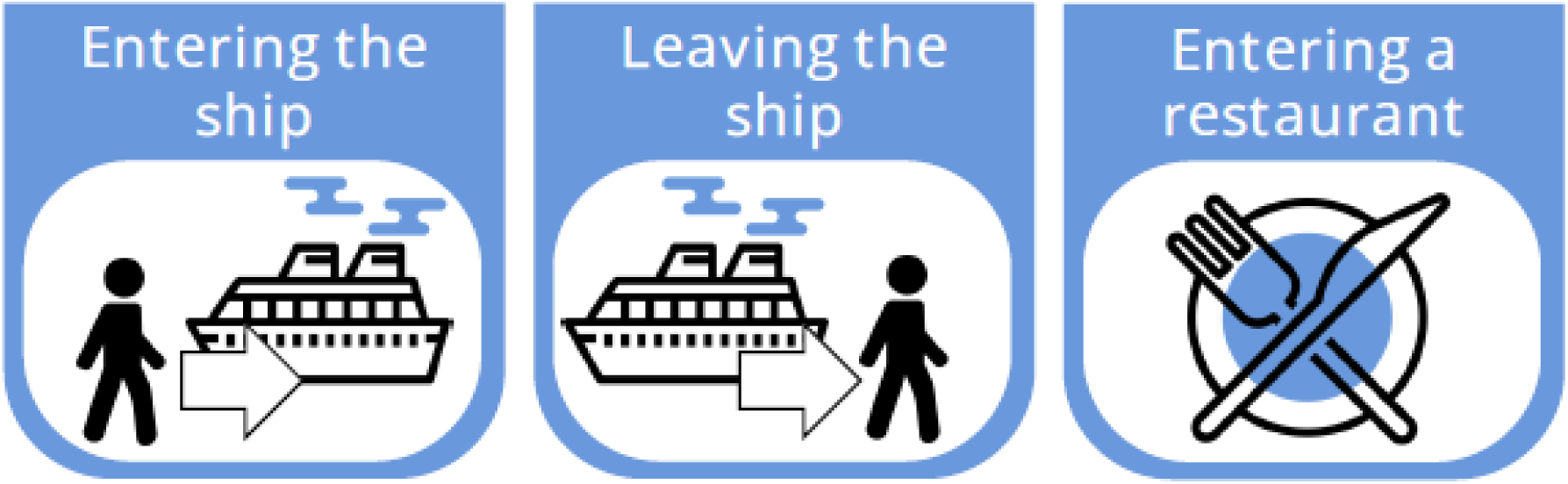
Hand hygiene moments that were monitored during the study.

Direct observation is considered the gold standard for measuring compliance. We recorded the number of people who performed hand hygiene and those who did not. The study focused exclusively on passengers, as crew members quickly recognized the observer, which could have introduced significant bias. Some of the crew may have been counted during embarkation or disembarkation if they were leaving the ship during their free time and wearing civilian clothes.

#### Questionnaire

An online questionnaire was created to assess the basic knowledge of crew members regarding infection control topics and to gather information about their educational background. The questionnaire included topics addressed during the training as well as general hygiene-related topics that were not directly covered. Some crew members completed the questionnaire before the training, while others did so afterward.

Responses to the general questions were used to measure the crew’s initial knowledge. This was particularly important since we were informed that the hand hygiene training could not exceed 30 minutes due to logistical constraints. Understanding their baseline knowledge helped us determine how to structure the training. Comparing the hand hygiene-related responses collected before and after the training made it possible to validate the effectiveness of our training.

Crew members were asked to fill out the questionnaire through their supervisors. A paper with a QR code linked to the questionnaire was provided to the supervisors. During their regular daily meetings, the supervisor showed the paper to the group of crew members, who scanned the QR code with their phones and completed the online questionnaire. The questionnaire is available in Supplementary Table 2.

#### The cruise ship

Sample collection was carried out aboard the *Celestyal Olympia* (Figure 5), a middle-size cruise ship sailing across the Aegean Sea. The vessel accommodates up to 1’664 passengers and 540 crew members. The ship has 12 decks, 2 large restaurants as well as 5 smaller bars. It is equipped with seven elevators and two main staircases.

**Figure 5:**
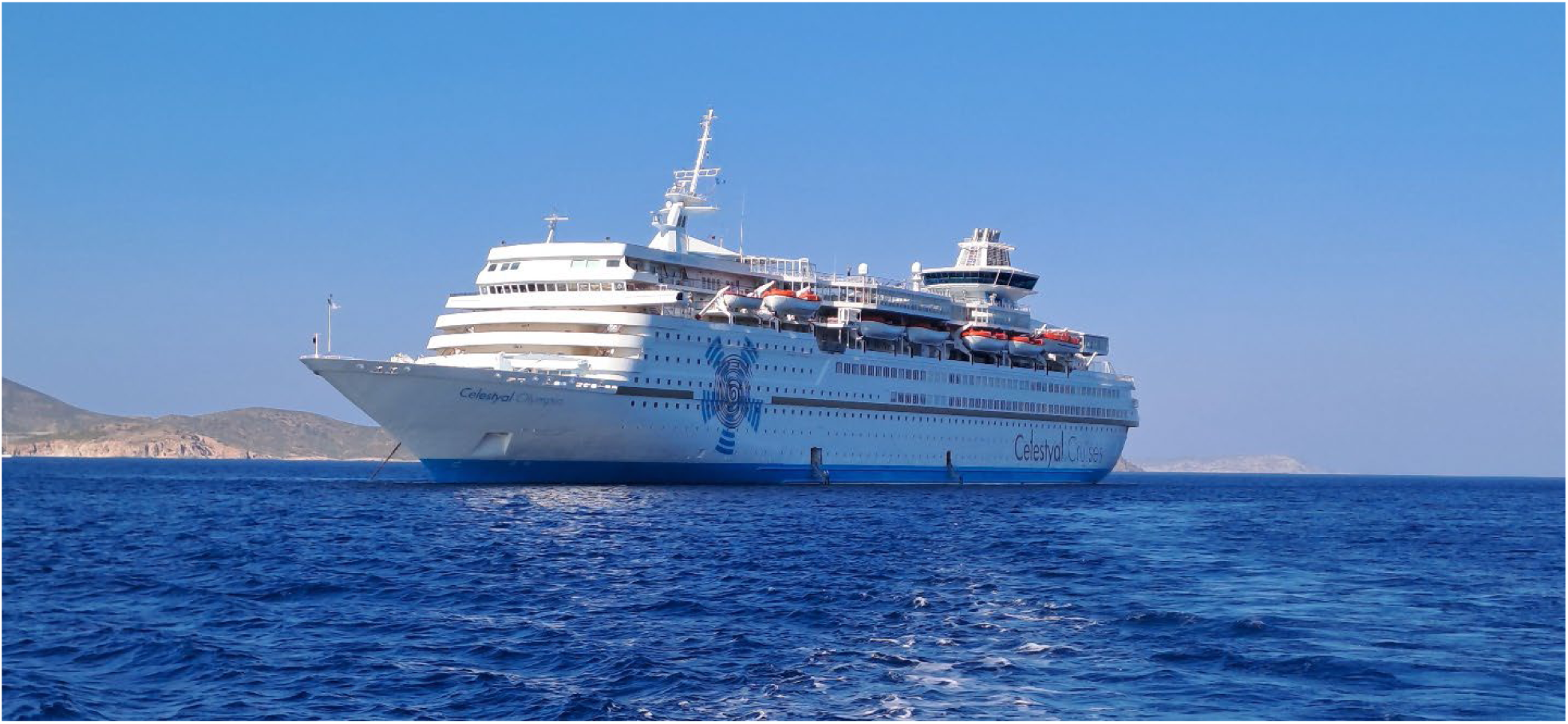
The cruise ship *Celestyal Olympia*, where sample collection was conducted.

#### Ethical Approval

As the project coordinator (University of Thessaly) is based in Greece and the work package leader (University of South-Eastern Norway) is Norwegian, the research proposal was submitted to both the Greek and Norwegian ethical boards. The Research Ethics Committee of the University of Thessaly approved the project (decision number: 59/19.09.2022). The proposal was submitted to the Norwegian Medical Research Ethics Committee (REK), which determined that the project falls outside the scope of the Health Research Act (i.e., it is not considered a clinical study) and therefore does not require ethical approval (reference number: 562942). The Norwegian Agency for Shared Services in Education and Research (Sikt) was notified of the project and granted approval (reference number: 164497).

#### Informed Consent

All participants provided written informed consent before taking part in the study. The consent process ensured that participants were fully informed about the study’s purpose, procedures, potential risks, and their right to withdraw at any time without consequences.

#### Data analysis

All data were stored and subjected to further analysis using Microsoft Excel (Version 2308, Build 16.0.16731.20310). A two-tailed paired t-test was used to compare microbial loads in Arm 2–4 to baseline. Fisher’s exact test was applied to calculate the one-tailed p-value to compare results collected before and after the training.

## RESULTS

### Hand hygiene product consumption

Unfortunately, we were unable to generate monthly statistics. The Provision Master’s log only recorded the hand hygiene products released from the main storage. However, after distribution to various units (e.g., housekeeping, restaurants), the products were stored in smaller supply rooms, and their usage was not documented. Additionally, the month-end closing dates differed between the two databases, making direct comparisons difficult. However, summarized data from a 15-month period can be analyzed together, and at this resolution, the data appears reliable.

Three types of hand hygiene product were used onboard: alcohol-based gel hand rub, regular soap, and antibacterial soap (Table 2). Antibacterial soap was primarily used in the kitchen, as it was odorless and tasteless. The next section details the number of night passengers and crew members spent onboard. Note that “months” do not necessarily align with calendar months; for example, August 2022 was recorded from July 22 to August 28. Product consumption per capita was calculated. For handrub gel and regular soap, total consumption was divided by the total number of people onboard (passengers and crew), as they all used the same products. Antibacterial soap, however, was used exclusively in kitchens and sanitization areas, where 79 crew members had access to it. Therefore, its consumption was calculated based only on this group.

**Table 2:**
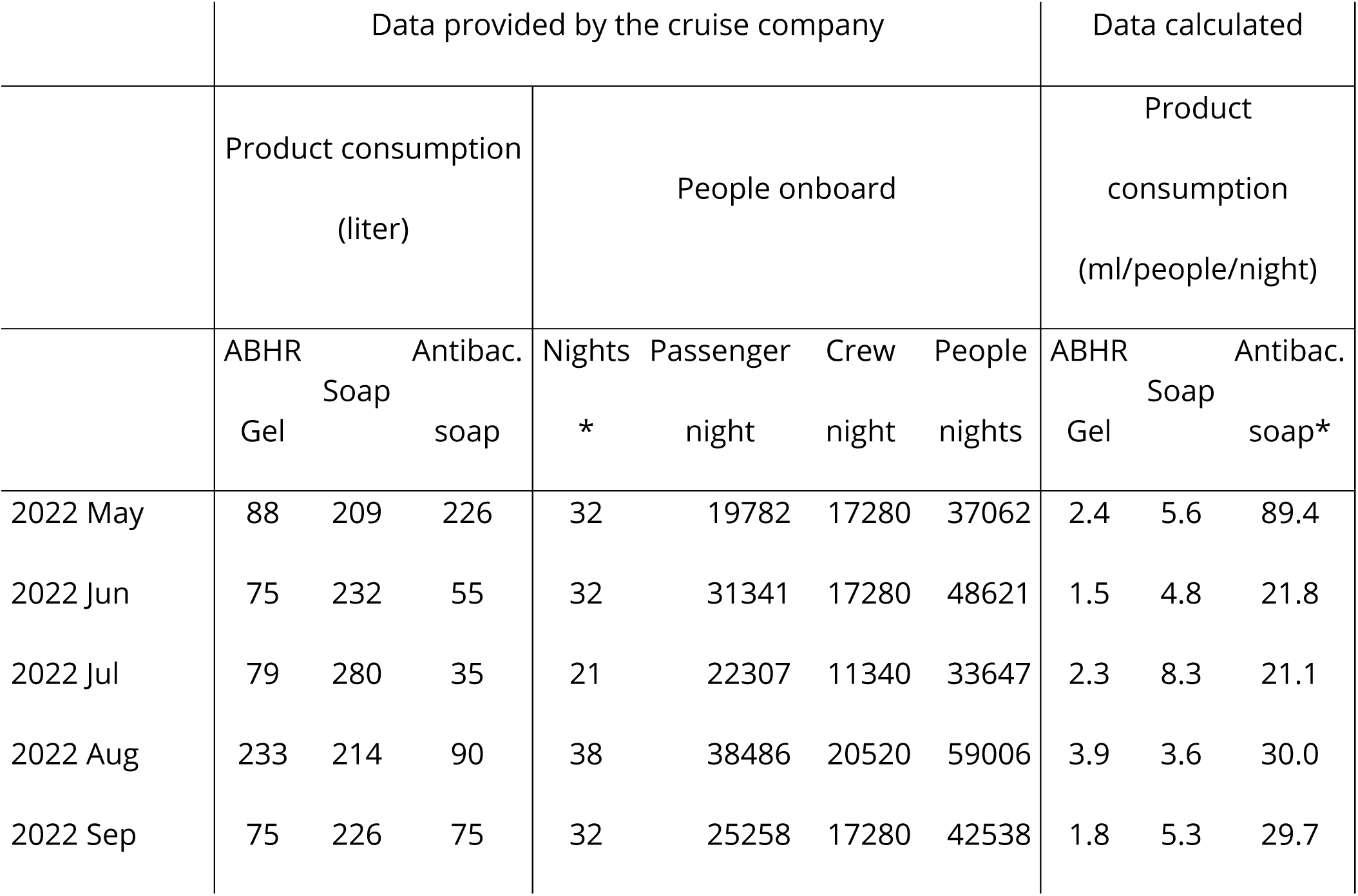
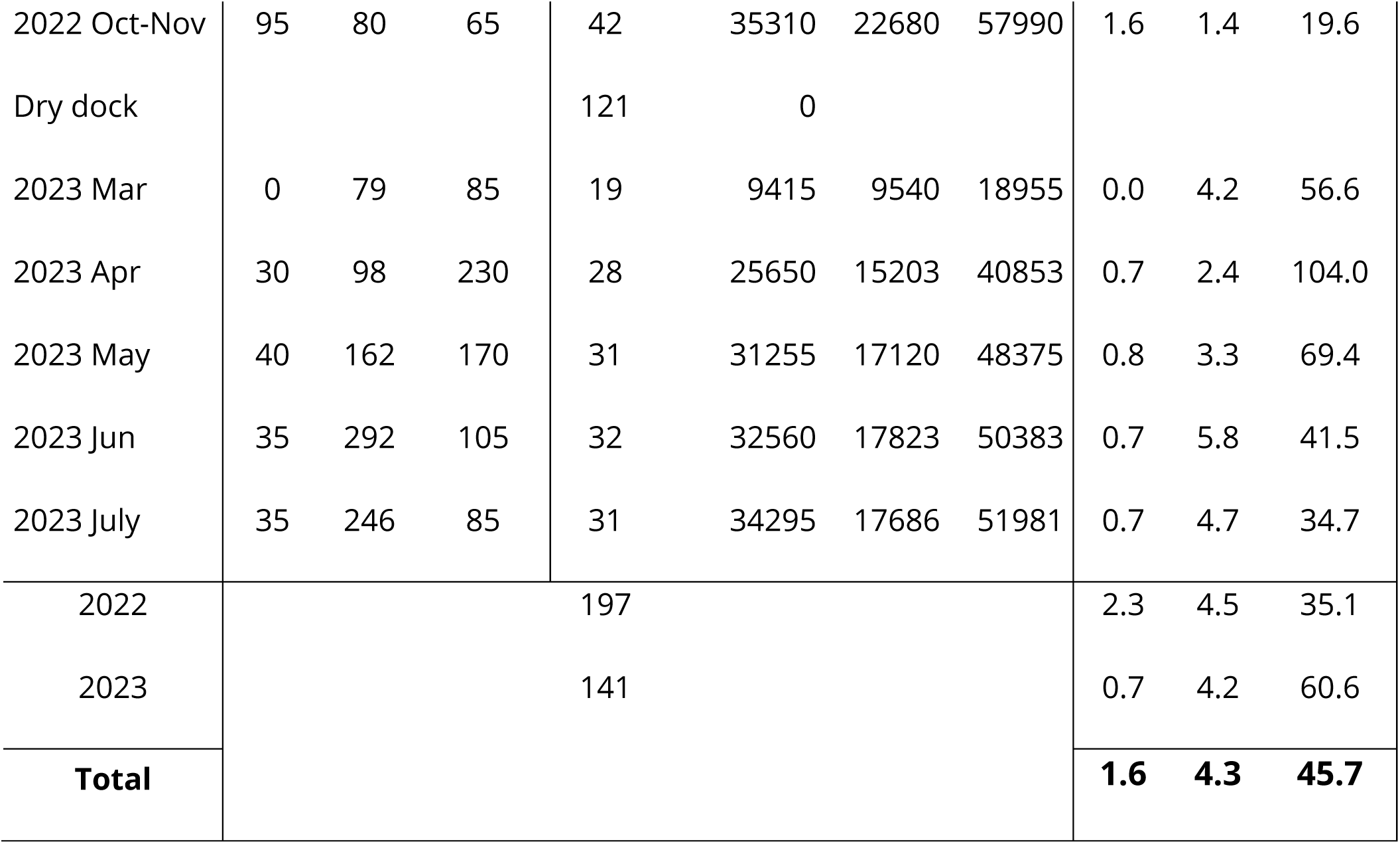
Hand Hygiene Product Consumption Onboard. *Note that months do not necessarily start on the first day or end on the last day of the calendar month.

Soap consumption remained relatively stable between the two years studied, whereas hand rub gel consumption significantly declined—from 2.3 ml/person/night in 2022 to 0.7 ml/person/night in 2023. This decrease can be attributed to the lingering impact of COVID-19 in 2022, when extra hygiene measures were still in place and heightened awareness of hand hygiene persisted. By 2023, conditions had returned closer to normal, making the lower consumption figure more representative of current trends. In contrast, antibacterial soap usage increased, suggesting that kitchen and sanitization staff were required to take hand hygiene more seriously.

On average, a person uses fewer than eight doses of soap per day and disinfects their hands less than twice daily (Table 3). Considering all situations where hand hygiene is necessary (e.g., after restroom use, before eating), this frequency appears insufficient. The issue is even more concerning when considering that 1 ml of gel handrub is often inadequate to cover the entire hand surface, double or even triple doses may be required for effective disinfection [6]. In contrast, antibacterial soap consumption is surprisingly high, indicating that kitchen staff take hand hygiene seriously; they perform hand hygiene frequently and use an adequate amount of product.

**Table 3:** Hand hygiene product consumption onboard together with dispenser output per dose.

|  | ABHR Gel | Soap | Antibac. soap |
| --- | --- | --- | --- |
| Product consumption (ml/people/day) | 1.6 | 4.3 | 45.7 |
| Average dose the dispenser provides (ml) | 1.0 | 0.6 | 0.7 |
| **Product consumption (dose/people/day)** | **1.6** | **7.6** | **65.3** |

**Table 4:** Compliance values and the number of observed hand hygiene moments during the four arms of the study.

|  | Hand hygiene compliance in %, (n) |  |  |  | n |
| --- | --- | --- | --- | --- | --- |
|  | Embarkation<br>(Entering the ship) | Disembarkation<br>(Leaving the ship) | Before entering the<br>Restaurant | Before entering the<br>Buffet | Total |
| Arm1 | 3.7% (2'408) | 1.8% (2'188) | 61.4% (1'120) | 4.7% (919) | 6'635 |
| Arm2 | 10.4% (1'555) | 7.0% (1'881) | 76.5% (885) | 9.1% (592) | 4'913 |
| Arm3 | 12.0% (1'505) | 23.5% (1'518) | 84.5% (889) | 10.7% (844) | 4'756 |
| Arm4 | 9.6% (2'058) | 6.3% (1'886) | 60.4% (866) | 6.5% (898) | 5'708 |
| Total | 8.9% (7'526) | 8.6% (7'473) | 70.2% (3'760) | 7,5% (3'253) | 22'012 |

**Table 4:** Statistical analysis of knowledge as an effect of the training. A significant shift was observed in only two cases.

| Question | Answer |  | Before<br>training | After<br>training | One-tailed<br>p-value |
| --- | --- | --- | --- | --- | --- |
| Handwashing is faster<br>than handrubbing | Correct | I'm sure it's false,<br>Probably false | 34 | 54 | 0.001* |
|  | Not correct | I'm sure it's true,<br>Probably true | 53 | 30 |  |
| Handwashing is more<br>skin-friendly than<br>handrubbing | Correct | I'm sure it's false,<br>Probably false | 7 | 32 | 0.000* |
|  | Not correct | I'm sure it's true,<br>Probably true | 80 | 52 |  |
| Rubbing your hands<br>with sanitizer usually<br>gets rid of germs<br>better than just<br>washing them with<br>soap and water. | Correct | I'm sure it's true,<br>Probably true | 63 | 72 | 0.064 |
|  | Not correct | I'm sure it's false,<br>Probably false | 22 | 13 |  |
| Handrubs are not<br>effective against some<br>germs | Correct | I'm sure it's true,<br>Probably true | 49 | 39 | 0.913 |
|  | Not correct | I'm sure it's false,<br>Probably false | 40 | 46 |  |

### Environmental microbiological samples

For environmental sampling, we aimed to select frequently touched surfaces. Samples were collected on three different occasions. One of these occasions was the lifeboat drill, mandatory for all passengers on the first day of the cruise. All master stations were located on Deck 7. People arrived gradually, but at the end of the drill, they all attempted to leave the open deck at the same time, creating a sharp increase in traffic (Figure 6). Samples #A and #B were collected immediately after this spike.

Figure 6: Traffic data during the mandatory lifeboat drill. Samples were collected from the door handle leading to the open deck and from the staircase rail.

*The original version of this figure included photographic materials that have been removed to comply with preprint policies prohibiting identifiable images. Readers interested in accessing the original visual data may contact the corresponding author for further information*.

This cruise ship visited two destination each day, giving passengers the opportunity to disembark twice daily for excursions. The morning disembarkation was more gradual, as the ship typically arrived at the port between 6:00 and 7:00 AM, but many passengers woke up later. In contrast, when the ship arrived to the afternoon destination, which occurred between 4:30 and 6:00 PM, nearly all passengers tended to leave at the same time. The crew made huge effort to organize this process, ensuring passengers could disembark as quickly as possible. Afternoon destinations included Mykonos, Patmos, and Santorini, where the ship was unable to dock directly at the port, requiring tender boats to transport passengers ashore. Despite the well-organized procedure, it often took up to an hour for all passengers to leave the ship. Figure 7 presents traffic data recorded during afternoon disembarkation. Three separate disembarkation events were analyzed: 1’053 passengers disembarked within an hour (Figure 7A). On average, 26.4% of them touched the stair rails (Figure 7B). Only 6% of passengers used the elevator during disembarkation (data not shown).

Figure 7: Traffic data during afternoon disembarkation. A: Number of people who used the left or right staircase. B: Number of people who used the left staircase, categorized by whether they touched the staircase rail. C: Staircase during disembarkation. D: Empty staircase. From this setting, samples were collected from the staircase rail, the interior and exterior elevator panels.

*The original version of this figure included photographic materials that have been removed to comply with preprint policies prohibiting identifiable images. Readers interested in accessing the original visual data may contact the corresponding author for further information*.

The third sampling occasion took place after the late-night show. During the night, Deck 5 hosted various entertainment programs. Passengers had the opportunity to watch the daily show at the Muses Lounge, enjoy drinks at the bar, or dance at the Selene Lounge (Figure 8B). As shown in the traffic data, there was a continuous, elevated flow of people for several hours during these events (Figure 8A). Samples were collected from restroom door knobs and staircase rails, both located between the Muses Lounge and the bar. Additionally, sample were taken from the door handle leading from the Selene Lounge to the open deck, the nearest designated smoking area.

Figure 8: Traffic data and sampling sites during late-night shows. The figure shows the number of people who exited Deck 5 using the left midship staircase (downward). Sample collection sites are marked with a red X on the floor plan.

Compared to the baseline arm, none of the intervention arms show significant differences in microbial load (Figure 9). There was no statistically significant reduction in bacterial counts on treated surfaces in Arm 2 compared to baseline (p > 0.3). Similarly, neither Arm 3 nor Arm 4 showed significant differences in surface CFUs from baseline.

**Figure 9:**
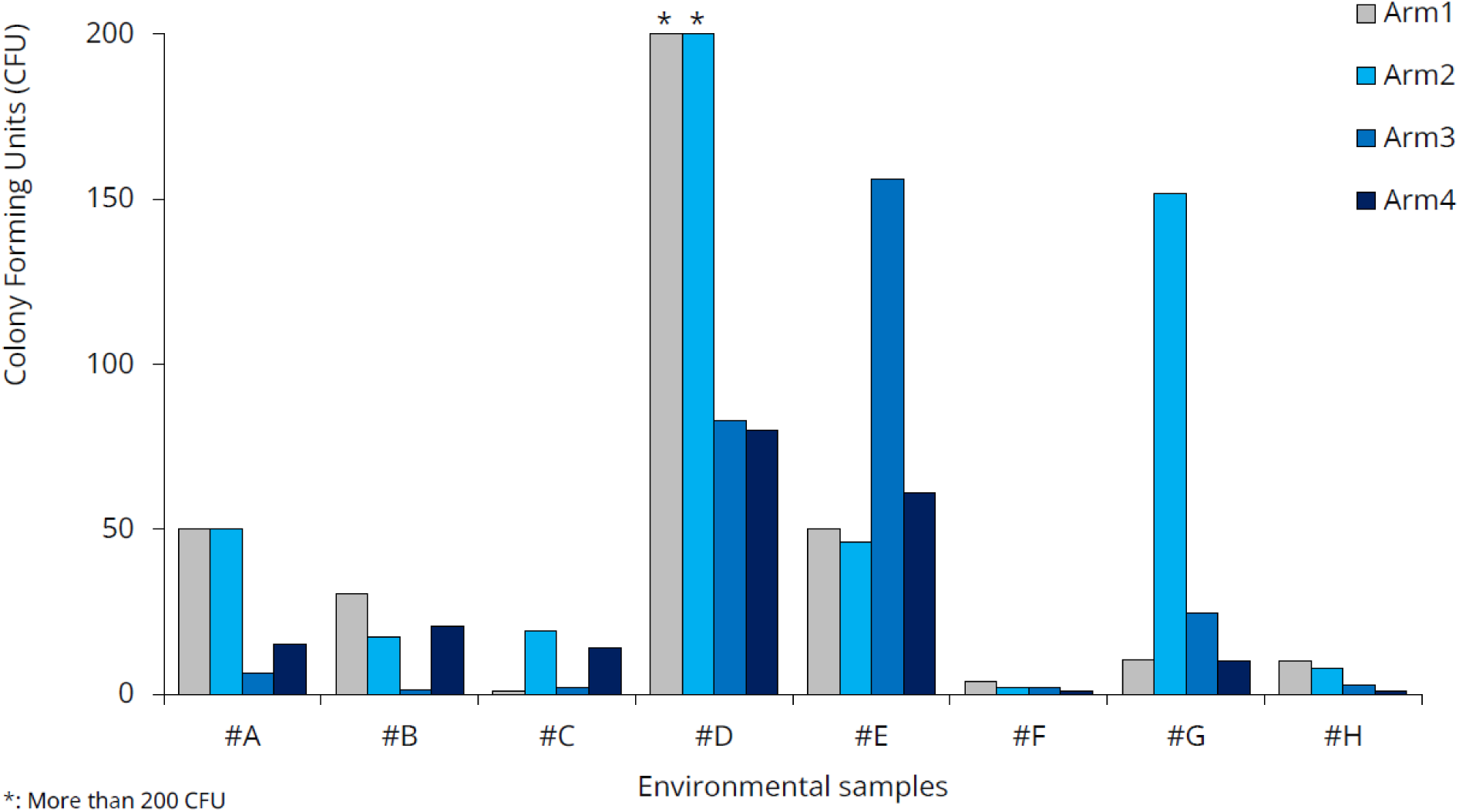
Colony Forming Units in the environmental samples.

### Hand hygiene compliance rate

22’012 hand hygiene opportunities were observed; however, we do not have data on the exact number of unique individuals these observations represent (Table 5, Figure 10). Due to the lack of passenger identification, some individuals may have been counted multiple times (e.g., once during embarkation and again when entering a restaurant).

**Figure 10:**
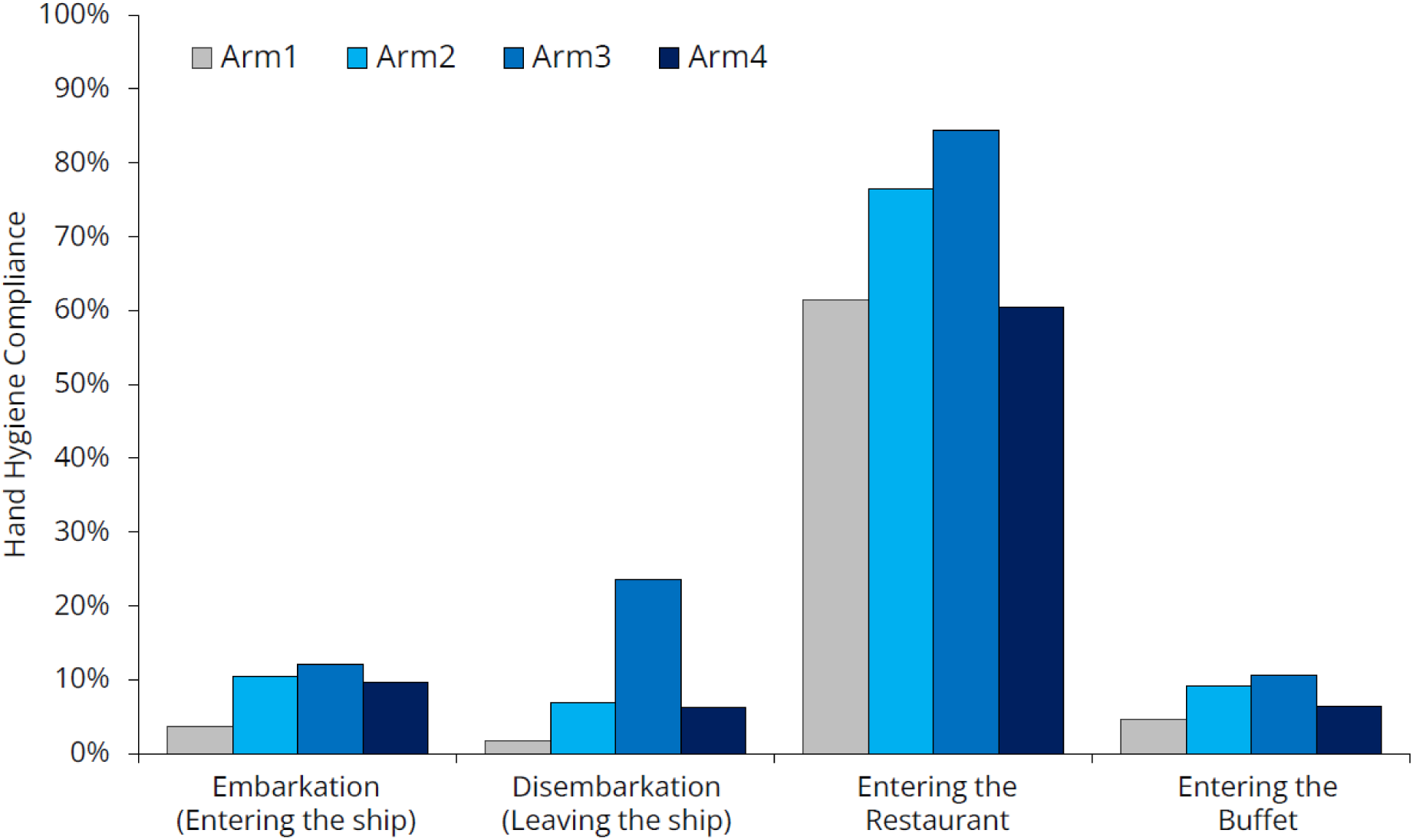
Hand hygiene compliance across different moments.

The most interesting observation during the study was that hand hygiene compliance was remarkably high before passengers entered the restaurant (70.2%) but remained low before they entered the buffet (7.5%). Passengers were free to choose whether to eat in the buffet or the restaurant, so the difference can only be partially explained by different passenger characteristics. The main difference between these two locations was that, at the restaurant entrance, a hostess not only asked how many people arrived together and helped them find a suitable table but also offered hand hygiene products (Figure 11). In addition to the dispenser, the hostess had a spray bottle to provide hand rub for everyone—a surprisingly cost-effective yet highly effective solution. At the buffet entrance, the same dispensers and spray bottle were left unattended, and no one reminded passengers to use them.

Figure 11: The main difference between the settings at the Restaurant (A) and Buffet (C) entrances. The hand hygiene product dispenser and spray bottle (B) were present in both cases, but only in the restaurant were they actively offered.

*The original version of this figure included photographic materials that have been removed to comply with preprint policies prohibiting identifiable images. Readers interested in accessing the original visual data may contact the corresponding author for further information*.

Note that even in the case of the restaurant, compliance never reached 100%. Most instances of noncompliance occurred when the hostesses were busy with a complex case and didn’t have the chance to engage with newly arrived guests. However, some passengers actively refused to perform hand hygiene. When the hostess was asked about the most common excuses, she reported that many passengers said rubbing their hands reminded them of COVID. It seems that some people became familiar with alcohol-based handrub during the pandemic and, as a result, perceive it as merely an emergency solution. Many people used the excuse, “I just had a shower.” Even the researcher overheard this excuse during compliance observations. Based on the intonation used by passengers, the message sounded more like, “I’m not that dirty that you need to disinfect me before letting me into the restaurant.” This suggests that these individuals do not fully understand the role of hands in infection transmission—or perhaps even the entire concept of hand hygiene. Addressing this issue is a far more complex challenge than what a hostess can resolve in just a few seconds.

Passengers also complained about the hand rub—both in general and regarding the specific product used onboard. Some refused to use hand rub, claiming it caused them skin problems. This concern can be valid for two reasons: if hadrub is used incorrectly, it can lead to skin issues (this topic will be discussed later). Additionally, during the pandemic, due to handrub shortages, anyone was allowed to produce handrub, some of which were of low quality because they lacked emollients. These low-quality products appear to caused long-term damage, leading some individuals to avoid handrub entirely. Other passengers stated that they generally accept handrubs but disliked the specific product available onboard. The handrub provided was a gel, which left a sticky feeling after use. Additionally, in some cases, residue remained on the hands as a whitish debris (Figure 12), which alarmed users who feared their skin was peeling due to the hand rub. Product acceptance is a crucial factor in improving compliance [7].

**Figure 12:**
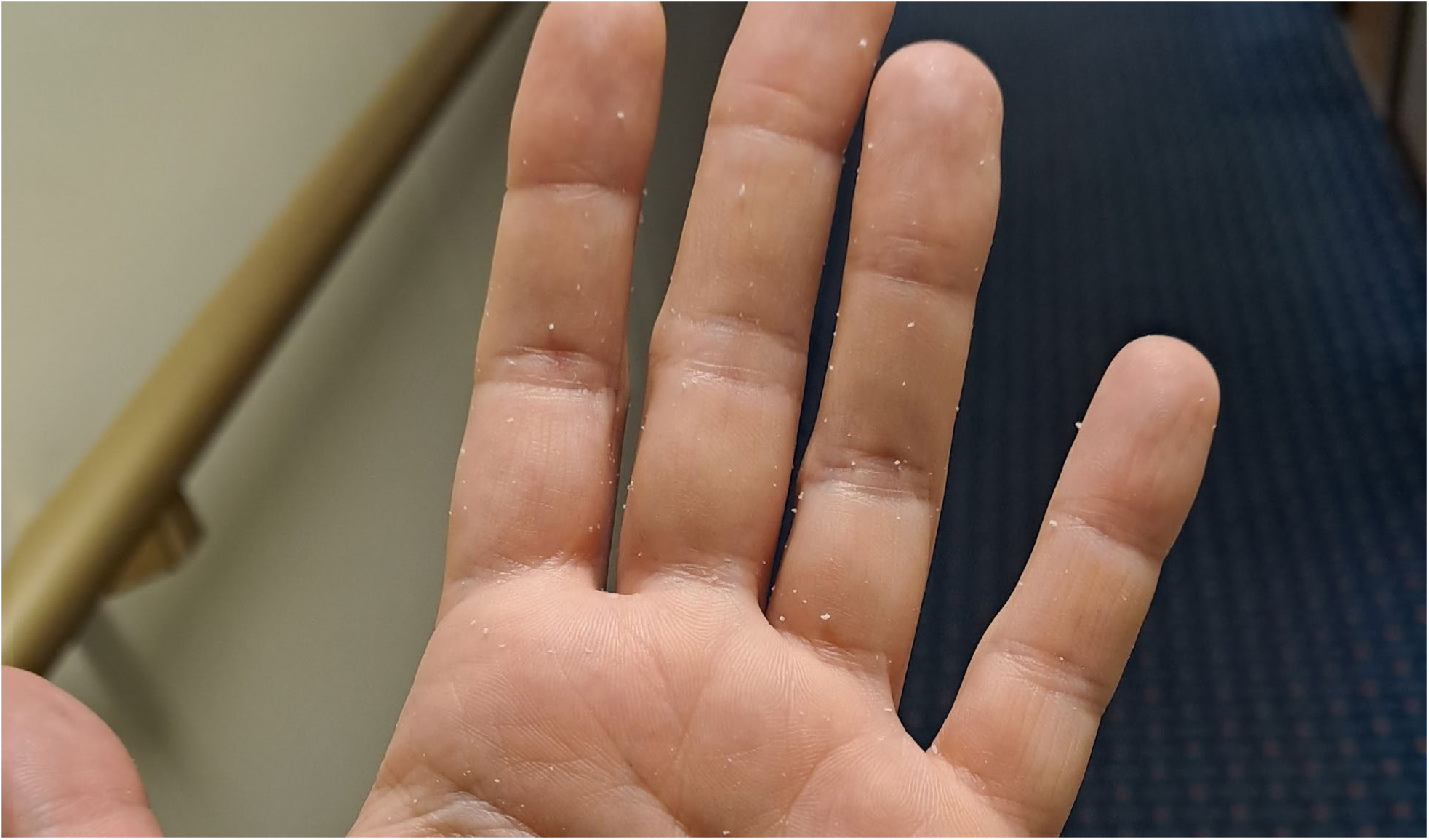
Residue of the gel handrub on the hand after hand hygiene.

The placement of the dispenser also had a major effect on compliance. We discussed this issue in a different communication [8]. Here, we highlight some settings in Figure 13 where compliance dropped to nearly zero. It is important to emphasize that the optimal location of the dispensers differs during embarkation and disembarkation, requiring repositioning of the dispensers (in our case, twice daily).

Figure 13: Examples of improper dispenser placement leading to a drop in compliance. A: The dispenser was placed in a corner because it was used to hold the rope. B: Dispensers were placed at the checkpoint where passengers had to present both their tickets and ship cards, leaving their hands full. C: The dispenser was hidden behind the door, out of sight of passengers returning to the ship.

*The original version of this figure included photographic materials that have been removed to comply with preprint policies prohibiting identifiable images. Readers interested in accessing the original visual data may contact the corresponding author for further information*.

During the observation, the researcher had an impression that young adults were less likely to perform hand hygiene, while elderly individuals and young children were more likely to do so. Elderly individuals are likely more concerned about health risks. Children have spent a significant portion of their lives during the epidemic, where hand hygiene was strictly enforced in nurseries and schools, making it a routine part of their daily habits. It was also observed that passengers from Asia and the United States performed hand hygiene more frequently, suggesting possible cultural differences in the acceptance of handrubs. Additionally, women tended to use handrub more often than men. In many instances, wife applied handrub first and then explicitly asked her husband to do the same. Future studies should explore these behavioral aspects further.

The placement of dispensers has a significant impact on compliance rates. The increase in compliance can be attributed to the fact that the researcher positioned the dispensers during Arm #2 and Arm #3, gradually identifying better locations. In contrast, the decline in compliance during Arm #4 can be explained by the fact that the researcher was occupied with crew training and was unable to closely monitor dispenser placement.

### Questionnaire

A total of 179 crew members completed the online questionnaire. After receiving the initial responses, we made slight modifications to the questionnaire. Regarding their educational background, the data suggests that crew were not undereducated, with most having attended school for 10 to 16 years (Figure 14). However, more than a quarter of them (n=50) reported never having studied biology.

**Figure 14:**
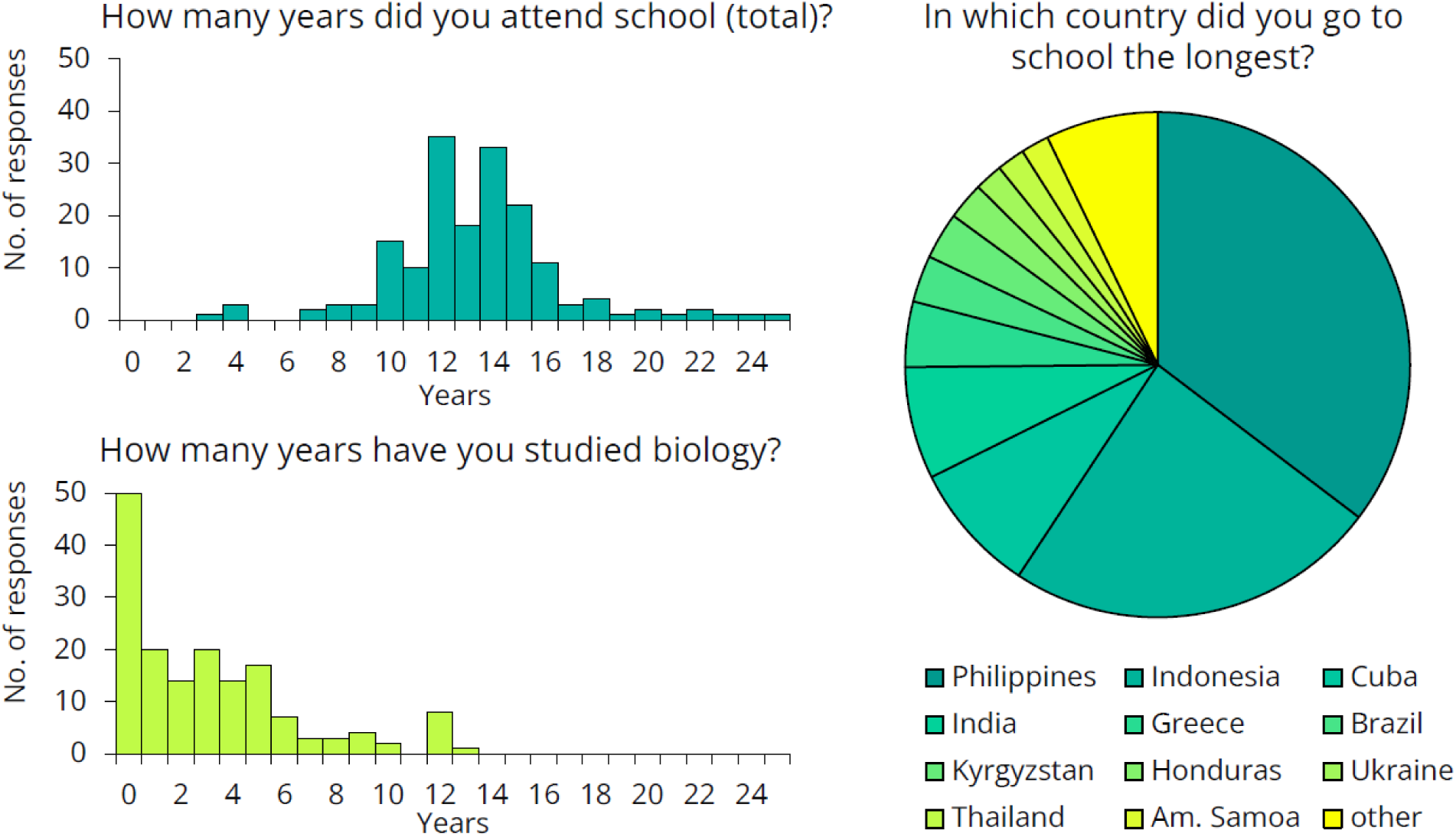
Crew members’ responses to the questionnaire on their educational background.

To better assess their preexisting knowledge, we presented several health-science-related statements and asked crew members to determine whether each statement was true or false (Figure 15). Only 69% of respondents knew that water is H₂O, suggesting that the training should avoid explaining the differences in chemical composition of disinfectants. Only one-third of them knew how many kidneys and livers humans have, suggesting that a training attempting to explain the exact workings of the immune system would likely not be effective. One-third of respondents were unaware that antibiotics kill bacteria or that not all bacteria are harmful. While 63% correctly recognized that bacteria existed on Earth before humans, 17% believed that dinosaurs were even more ancient. This suggests that complex topics such as the endosymbiotic theory, the bacterial origins of mitochondria, and their implications for antibiotic use should be excluded from the training. Twenty-three percent of the participants believed that bacteria do not have DNA, suggesting that it would be challenging to impress them with the latest sequencing results — that was performed as part of the same intervention study [15]. As we shifted from bacteria to viruses and yeast, the results worsened, with the responses becoming nearly evenly split, around 50-50%. The data strongly suggested that we needed to design the training in a way that did not rely on any preexisting knowledge.

**Figure 15:**
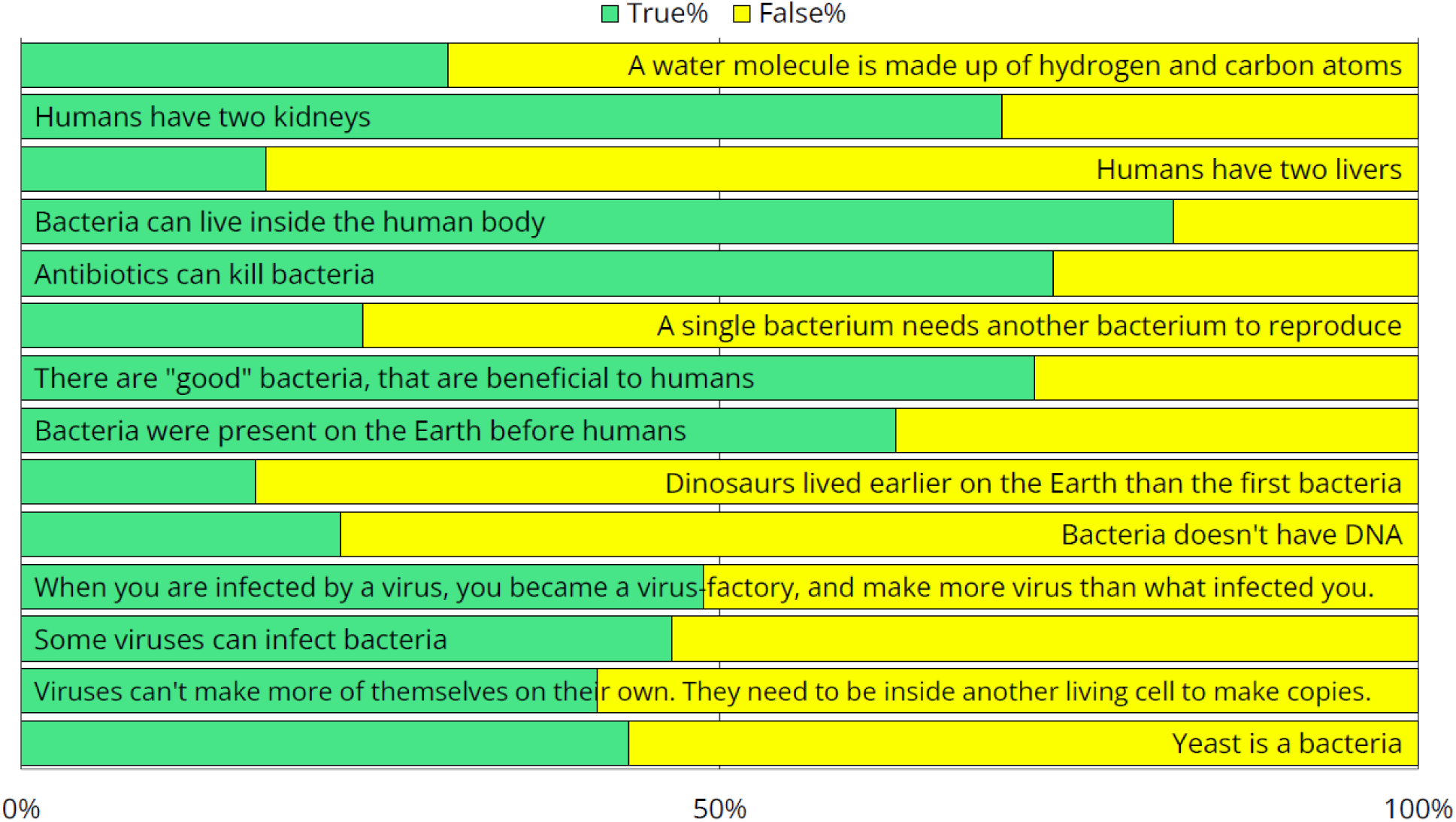
Responses to survey questions aimed at measuring preexisting health-science knowledge. Note that since these were yes-or-no questions, answers with a distribution near 50-50% indicate that the respondents had no clear idea.

### Interventions

#### Surface disinfection

As evident from the environmental sample data, there were no significant changes in bacterial load during the surface treatment arm (Arm 2) compared to the baseline arm (Figure 9). When analyzing the effects of the sprays separately, neither showed a significant difference compared to the baseline. The p-value was 0.082 for Spray #A and 0.336 for Spray #B.

#### Technology induced behavioral change

The Semmelweis System was installed onboard on September 1, 2023, and remained operational until November 9. During this period, a total of 1’636 measurements were conducted, with 204 QR codes in use. The system’s primary advantage is that it provides immediate feedback about missed surfaces, initiating a learning process.

The system should be used multiple times, as development occurs continuously during the first 5 to 20 uses [9,10]. Although more than 200 crew members tried the system, only 76 used it at least twice (Figure 16). Only 10 participants used it 10 times, which was the desired number for analyzing improvement. The results from these 10 participants are shown on the left. In their case, the system effectively taught the correct hand hygiene technique; the percentage of missed hand surfaces was initially 9.6%, which reduced to 1.6% by the 10th use.

**Figure 16:**
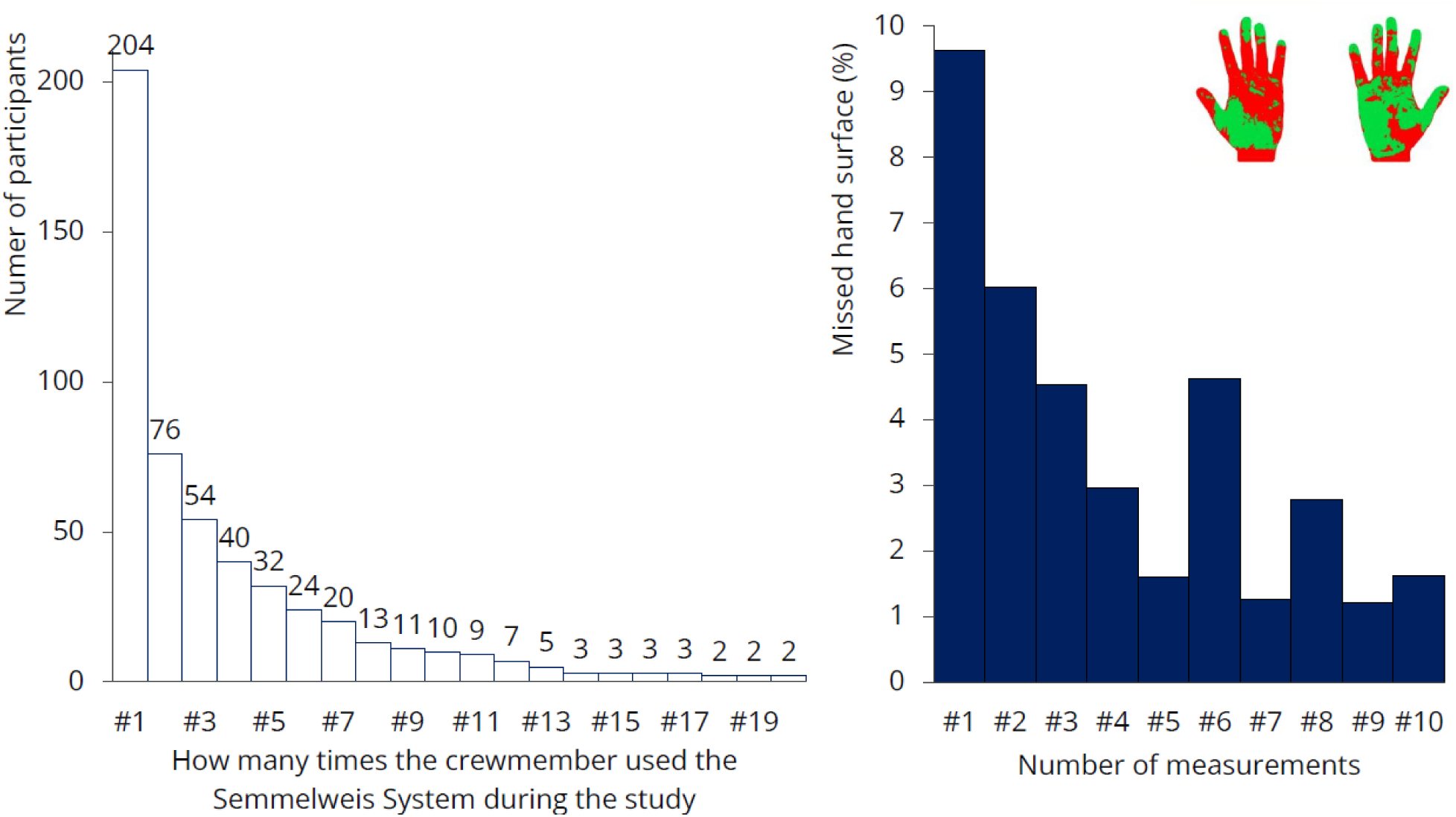
Results of the hand hygiene monitoring. Left: the number of times participants tested themselves is shown. Only 10 participants used the system 10 times. Right: results of that 10 people: missed hand hygiene surfaces decreased from 9.6% to 1.6%.

Unfortunately, teaching 10 crew members how to properly rub their hands will not bring significant change on a ship with 540 crew members. The researchers asked the some of the crew why they were not using the system more frequently. They mentioned that they were extremely busy throughout the day and hand hygiene was simply not a priority for them — at least not enough to spend their free time practicing it. The system was used most frequently by restaurant staff, where managers were highly committed to hand hygiene and encouraged the crew to perform the test during their shifts. A possible solution might be to make the use of such monitoring devices mandatory for the crew while on duty. These measurements will not be effective if crew members are required to do it during their free time, just for fun.

#### Hand Hygiene Training

The passengers’ pillow letter was ineffective (Figure 9). Despite the fact that the surveyed moments were clearly highlighted in the flyer, compliance decreased in Arm4 compared to the previous arm. While we don’t have a clear explanation for this, some phenomena were observed. Language barrier was evident. Although the official ship language was English, many passengers arrived in groups with a group leader who translated for them, meaning they did not necessarily understand English. A one-language flyer may not be effective.

The crew’s hand hygiene training was conducted in small groups and structured as an interactive discussion rather than a traditional lecture. The content was continuously refined based on feedback collected by the researcher. The latest version of the training material can be found at the following link: https://sites.google.com/view/usn-hs-2023/traning-material. We would be happy to offer this version as a starting point for further development of training materials.

During the training, some crew members had the opportunity to participate in an experiment (Figure 17). Volunteers were asked to touch the left side of a contact plate with a finger. They then performed hand hygiene using the handrub gel commonly used onboard and touched the right side of the plate with their disinfected finger. After culturing, the plates were photographed, printed, and displayed in the corridor next to the crew mess, where crew members eat three times a day. Plates from the previous day’s experiments were also presented during the training session.

**Figure 17:**
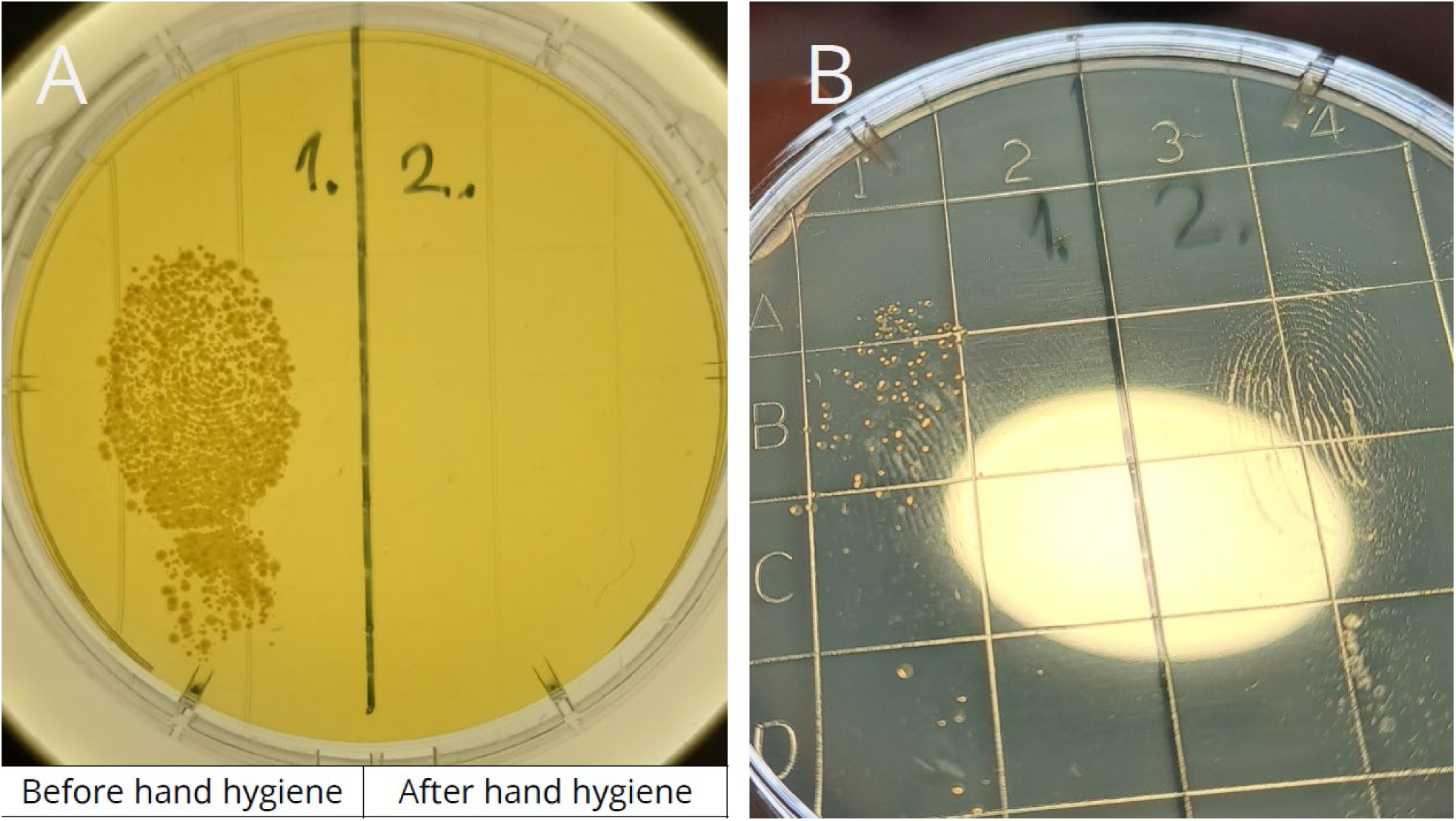
The hand hygiene experiment demonstrates the effectiveness of hand hygiene. During crew training, volunteers pressed one finger onto the left side of a contact plate before hand hygiene and onto the right side after disinfection.

Based on 19 experiments, the average colony-forming unit (CFU) count before hand hygiene was 150.1, which decreased to 1.5 after hand hygiene—representing a log_10_1.99 reduction. However, this reduction is likely underestimated, as colonies in the “before” samples were often so dense that they merged, making precise counting difficult.

Some of the crew members attended the hand hygiene training before (n=79) and some after (n=69) filling out the questionnaire. The questionnaire included questions comparing soap-and-water handwashing with hand rubbing using alcohol-based handrub. Since this topic was directly addressed during the training, comparing the knowledge of the “before” and “after” groups provides a valuable opportunity to assess the effectiveness of the training.

In all four questions, there was some shift, with more participants answering correctly after the training than before (Figure 18, correct answers marked by a star). We were not fully satisfied with the extent of this improvement, indicating that there is still room for improvement how the message was targeted (Table 1).

**Figure 18:**
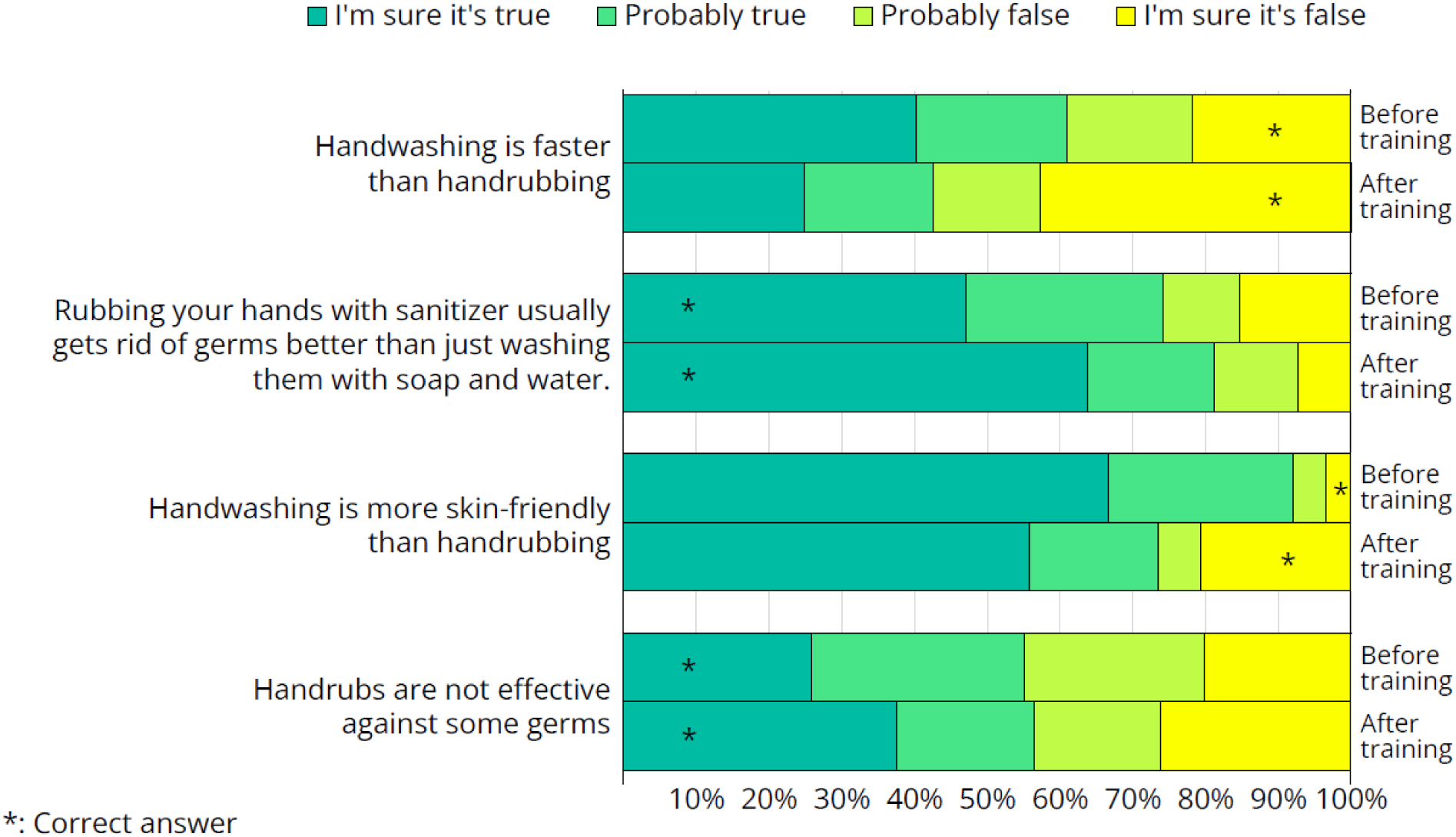
Responses from the questionnaire on hand hygiene-related questions that were directly addressed during the training. Some crew members completed the questionnaire before the hand hygiene training, while others did so afterward.

Less than half of the crew members were aware that hand rubbing takes less time than hand washing, despite this being one of the primary reasons alcohol-based hand rubs were introduced—first in healthcare and later in other sectors. Proper hand washing takes approximately 1.5 minutes, whereas handrubbing can be completed in just 30 seconds [11].

Alcohol-based handrubs are more effective at removing microorganisms than soap-and-water hand washing [7, 12]. Alcohols exhibit excellent in vitro germicidal activity against vegetative bacteria and fungi. However, they have virtually no activity against bacterial spores or protozoan oocysts and are less effective against some non-enveloped viruses. At first glance, this may seem contradictory, making it difficult for participants to understand how handrubs can be more effective overall if they are ineffective against certain germs. We believe this is a critical concept, and future training sessions should dedicate more time to explaining it clearly.

Alcohol-based solutions or gels containing humectants cause significantly less skin irritation and dryness than soap [7]. This was surprisingly new information for many crew members. During the training, several participants expressed skepticism. They mentioned that their skin problems started when hand rubs were introduced onboard. Upon further discussion about how exactly they are using handrub, it became evident that many crew members were applying to gel immediately after handwashing, while their hands were still wet. The World Health Organization’s Hand Hygiene Guidelines explicitly state that washing hands with soap and water immediately before or after using an alcohol-based product is not only unnecessary but may also lead to dermatitis [7]. The European ShipSan Manual for Hygiene Standards on Passenger Ships suggests that sanitizer may be used after washing, but it also defines proper handwashing as rubbing the hands with soap in hot water, followed by drying [13]. The CDC Vessel Sanitation Program Manual states that hand antiseptics should be applied to clean hands but does not explicitly mention the need to dry hands first [2]. However, it does specify that soap, paper towels or air dryers, and a waste towel receptacle must be available at handwashing stations, implying that drying is necessary. Future training sessions should emphasize that handrub should never be applied to wet hands.

Alcohol-based solutions containing 60%–95% alcohol are the most effective [12]. Applying hand rub to wet hands dilutes the alcohol content, which not only increases the risk of skin irritation but may also reduce its effectiveness. In addition, damaged skin may harbor more pathogens than healthy skin [14]. Future training sessions should emphasize that hand rub should never be applied to wet hands.

## DISCUSSION

Microbial loads of frequently touched surfaces have not changed significantly; none of the employed interventions were able to reduce bacterial load. Environmental microbiological samples only measured the CFU count. The microbial composition of these samples was analyzed in a separate publication [15], where the sample codes were slightly different: #A = #A1, #B = #A2, #C = A5, #D = A6, #E = #A4, #F = #A3, #G = #A10, and #H = #A8.

The location of the dispensers largely influenced hand hygiene compliance. In a previous communication [8], we outlined key recommendations for optimal dispenser placement to maximize effectiveness: dispensers should be clearly visible from a distance, allowing people time to process and react. They should be placed in areas where people walk slowly and are not in a hurry. Dispensers should be positioned along natural walking paths, as people are unlikely to take extra steps to reach them. People should have free hands when approaching a dispenser. When required to show cards, tickets, their hands are not free, so they cannot perform hand hygiene. When they hold items, many attempted hand hygiene by dispensing sanitizer onto only one hand or the back of their hands, creating an illusion of proper hand hygiene without achieving effective bacterial reduction. Some people holding items (ship cards, phones, wallets) in their mouths to free their hands, which, from an infection control perspective, is an even worse alternative than skipping hand hygiene altogether. The high compliance observed at restaurant entrances clearly indicates that reminders play a crucial role in encouraging dispenser use.

Compliance at the “before entering the restaurant” moment dropped from 84.5% in Arm 3 to 60.4% in Arm 4. According to the hostess, an incident occurred during the gap between the study arms: she offered handrub to a passenger who reacted aggressively, yelling at her and throwing the sanitizer onto her clothes and into her eyes. The situation was so extreme that she broke down in tears in front of everyone. Since then, she reported that she felt uncomfortable offering handrub to passengers. This incident serves as a critical reminder that cruise companies must prioritize the motivation and mental well-being of their staff. A single traumatic event led to a 15% drop in compliance in the following weeks, potentially increasing the infection risk for thousands of passengers.

In a previous study, four antimicrobial surface treatment sprays were tested, two of which were the same as those used in this study. That study was able to validate their effectiveness *in vitro* when bacteria were applied in suspension. However, in real-life settings, the sprays did not demonstrate the same level of efficacy [16].

Regarding hand hygiene training, based on the feedback collected, several general conclusions can be drawn. The training cannot assume any prior infection control-related knowledge. Questionnaire results indicate that while crew members are not undereducated, their education often lacks a background in biology. Additionally, a language barrier exists in this area. Although crew members speak English well enough for their daily tasks, biological terminology is not necessarily part of their vocabulary. To address this, we recommend that the training rely heavily on images and videos rather than text. If feasible, translating the training materials could further enhance comprehension and engagement.

It should be mentioned, that we encountered various conspiracy theories among crew members, mostly regarding COVID-19, but also vaccination, mask-wearing, disinfection, and other various health-related topics. The hand hygiene experiment was especially effective, as it allowed crew members to witness the results firsthand rather than simply being told what to believe. Observing the outcomes of their own experiments from the previous day made the lesson more tangible and credible.

Additionally, crew members were surprised by the effectiveness of their hand rub. Many expressed dissatisfaction with the handrub used onboard, describing it as “sticky” and uncomfortable, as it was previously mentioned. This led them to associate the product with low quality and to doubt its germicidal efficacy.

### Limitations

This study provides valuable insights into hand hygiene interventions on cruise ships; however, several limitations should be considered when interpreting the findings.

### Study Design Limitations

The study employed a sequential intervention design on a single cruise ship without a parallel control group. While each intervention arm was compared to the baseline, the lack of a concurrent control means that external factors, such as increased general awareness over time or study fatigue, may have influenced compliance rates. For example, by the time the hand hygiene training was introduced, passengers and crew may have already become more familiar with the study, potentially affecting their behavior independently of the intervention itself. Similarly, situational factors, such as the observed negative passenger-staff interaction affecting compliance enforcement, could not be isolated from intervention effects. Despite these challenges, the use of a defined baseline arm provides a reasonable control for assessing changes across study arms, and the ship’s operational environment remained relatively stable during the study period, allowing for meaningful comparisons. Future studies employing a randomized or multi-ship design could further strengthen causal inferences.

### Sample Size and Data Limitations

Certain aspects of the study had limited sample sizes, which may impact the generalizability of the findings. The environmental microbiological sampling included only eight surfaces, which, while representative of high-touch areas, may not capture broader microbial trends across the ship. Additionally, engagement with the hand hygiene monitoring device was lower than expected; although 204 QR codes were issued, only 10 crew members completed the recommended 10 scans necessary for meaningful assessment of hand hygiene improvement. This limited data prevents definitive conclusions on the long-term effectiveness of the device-based intervention.

Hand hygiene compliance observations, while extensive in total (over 22,000 opportunities recorded), did not track individuals over time. Thus, the dataset reflects overall compliance rates at different locations rather than behavioral changes in specific passengers. As a result, it is unknown whether the same individuals improved their compliance or whether compliance levels fluctuated among different groups of passengers. The crew questionnaire was also subject to selection bias, as responses were voluntary and self-reported, meaning those with a stronger interest in hygiene may have been overrepresented. While these factors do not invalidate the findings, they should be considered when applying the results to broader cruise ship populations.

### Potential Biases

The presence of observers recording compliance may have introduced the Hawthorne effect, wherein passengers modified their behavior due to awareness of being watched. While observers were instructed to remain discreet and dressed in plain clothing to minimize this effect, some passengers may have still noticed them. This could lead to an overestimation of compliance rates compared to unobserved, real-world conditions. Additionally, the crew’s participation in the questionnaire and training may have been influenced by social desirability bias, where respondents provided answers they perceived as favorable rather than reflecting their actual knowledge or behavior.

Despite these limitations, the study offers valuable practical insights into the challenges and opportunities for improving hand hygiene on cruise ships. Future research with larger, randomized samples and more controlled conditions could further refine these findings and contribute to the development of stronger, evidence-based guidelines for infection control in maritime settings.

## CONCLUSIONS

Hand hygiene product consumption data suggests that hand hygiene practices onboard need improvement, with the exception of the kitchen area, where standards appear to be well maintained. Notably, during our study, when we mentioned that we were there to improve hygiene, most people immediately associated it with food hygiene. This indicates that food hygiene is a well-recognized area with clear guidelines and regular monitoring, leading to high standards. In contrast, other areas, such as passenger hand hygiene, receive less attention and have fewer established recommendations. This also suggests that cruise companies tend to comply with hygiene guidelines and regulations when they exist, highlighting the need for clearer recommendations in understudied areas like passenger hand hygiene.

From the compliance data, we can see that some people are aware of the importance of hand hygiene and perform it under any circumstances (they carry their own handrub or actively seek out dispensers), but this represents less than 5% of the passengers. There is also a small group, less than 10%, who are strongly committed to NOT performing hand hygiene, for whatever reason. For the majority of passengers, hand hygiene is simply not a priority. If they find a dispenser in the right place at the right time, or if they are directly asked to use it, they will comply. However, they will not make any extra effort to do so. This highlights the importance of establishing clear recommendations for cruise ship companies on how to create an effective hand hygiene setup.

Our study also underscores that, unfortunately, there is no simple solution to improving hand hygiene. Measures such as spraying surfaces once a month, placing informational leaflets in cabins, or installing an unattended control machine in the hall are unlikely to significantly change hand hygiene behavior on their own. Hand hygiene interventions need to be more complex, and it appears that human resources will be essential—crew members should be specifically assigned to promote and facilitate hand hygiene.

These findings can inform cruise operators and public health guidelines – emphasizing convenient access to hand hygiene, active promotion by crew, and selecting acceptable products could collectively raise hygiene standards and reduce outbreak risks on ships.

## Data Availability

All relevant data are within the manuscript and its Supporting Information files.

## ACKNOWLEDGEMENTS

HEALTHY SAILING project has received funding from the European Union’s Horizon Europe Framework Programme (HORIZON) under Grant Agreement number 101069764. Funded by the European Union. Views and opinions expressed are however those of the author(s) only and do not necessarily reflect those of the European Union or the European Climate, Infrastructure and Environment Executive Agency (CINEA). Neither the European Union nor the granting authority can be held responsible for them. This work was funded by UK Research and Innovation (UKRI) under the UK government’s Horizon Europe funding guarantee [grant number 10040786], [grant number 10040720]. This work has received funding from the Swiss State Secretariat for Education, Research and Innovation (SERI). We thank all crew members of *Celestial Olympia* for supporting the study.

## AUTHOR CONTRIBUTIONS

SB: Methodology, Investigation, Writing – Original Draft Preparation HS: Project Administration, JK: Conceptualization, Supervision, Writing – Review & Editing

---

**Supplementary Table 1:**
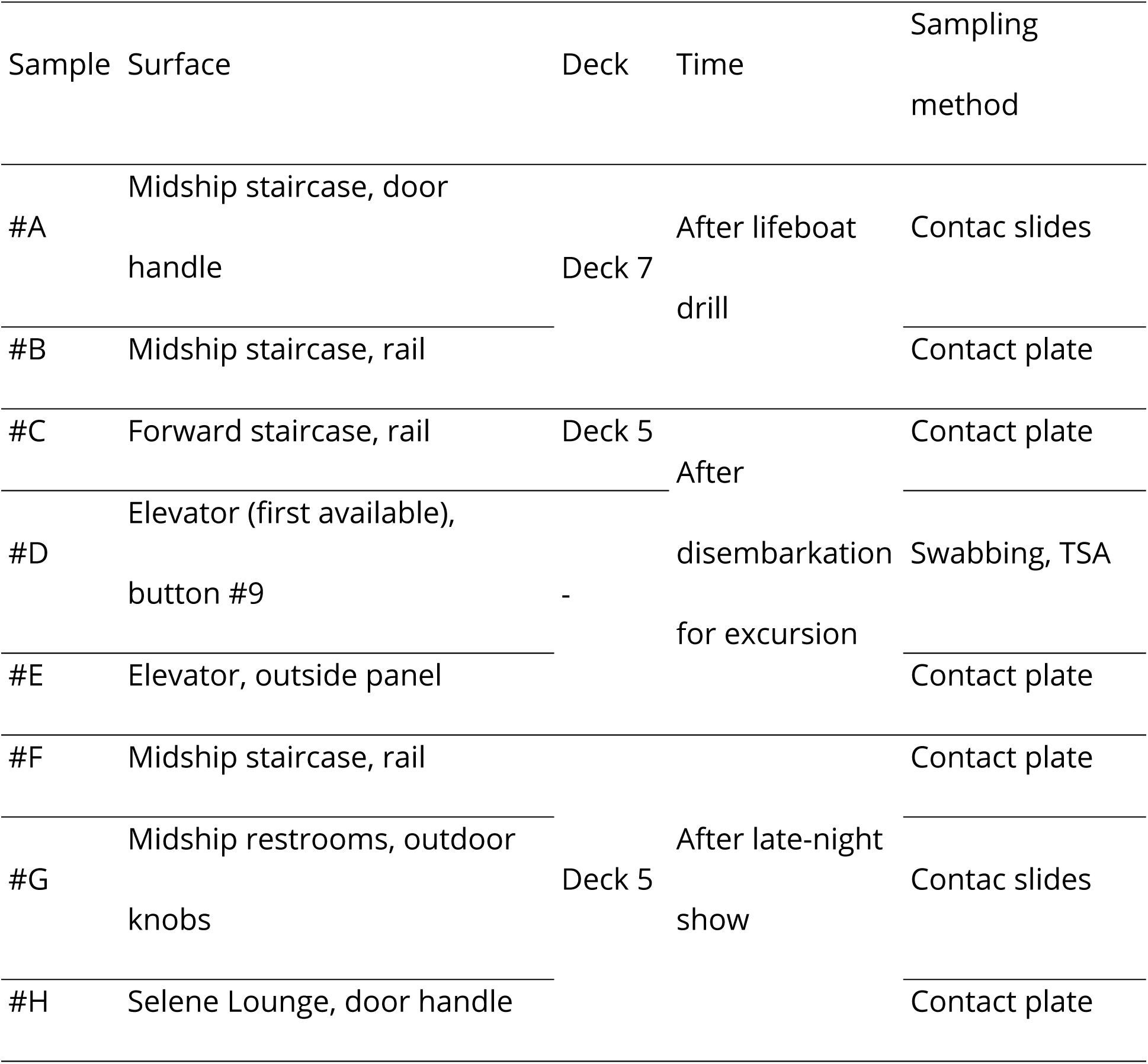
Description of the sampled surfaces and sample collection methods.

**Supplementary Table 2:**
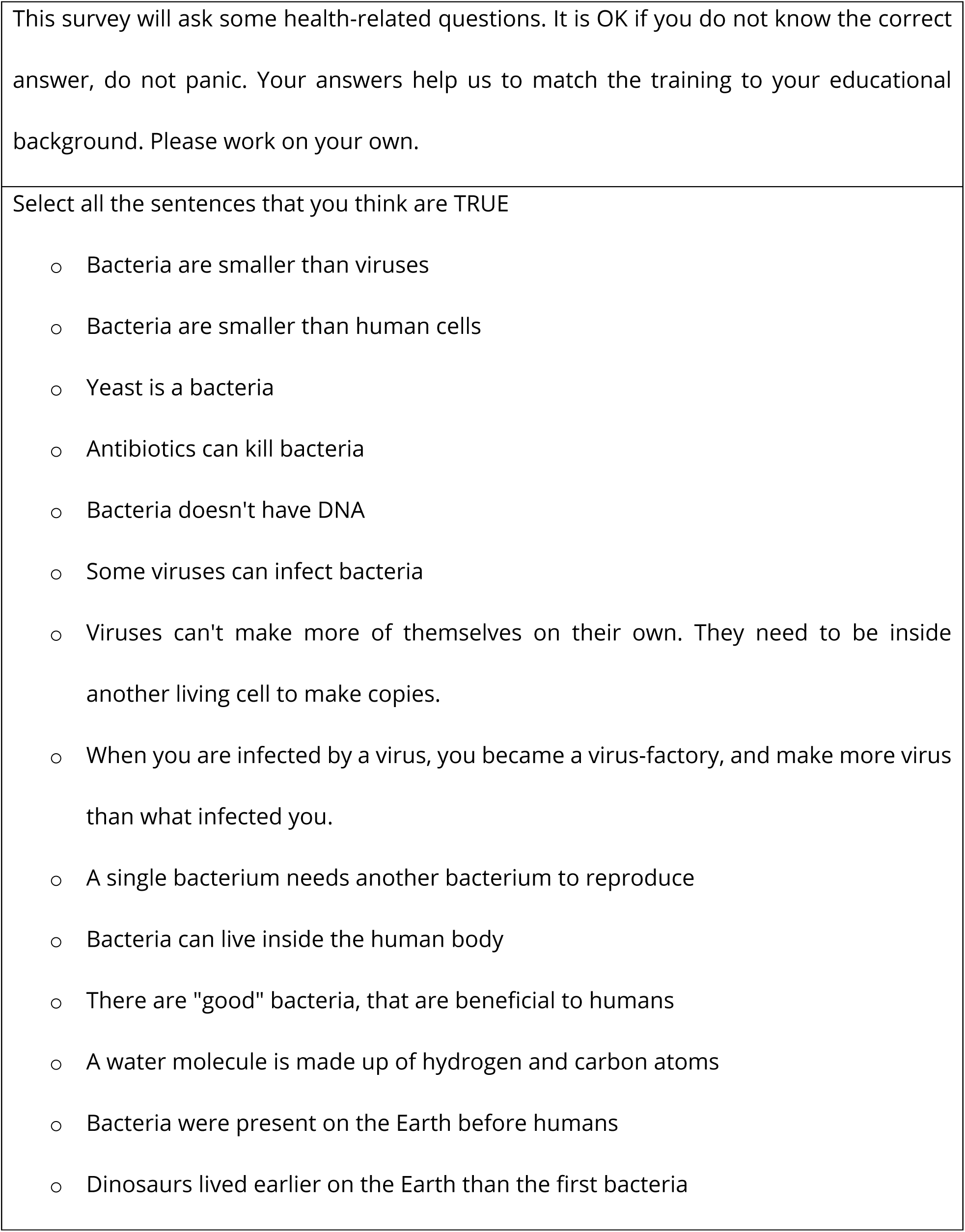
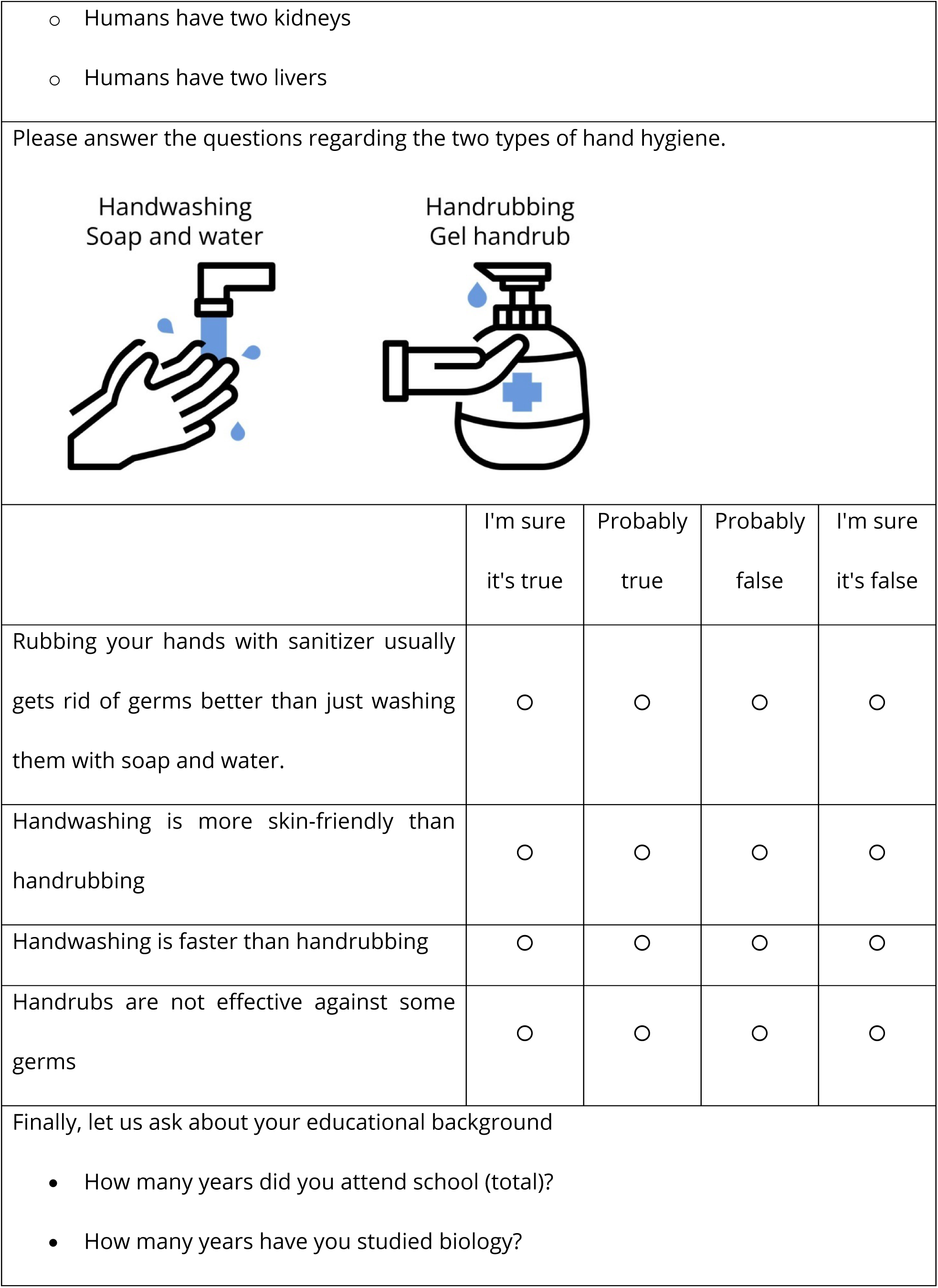
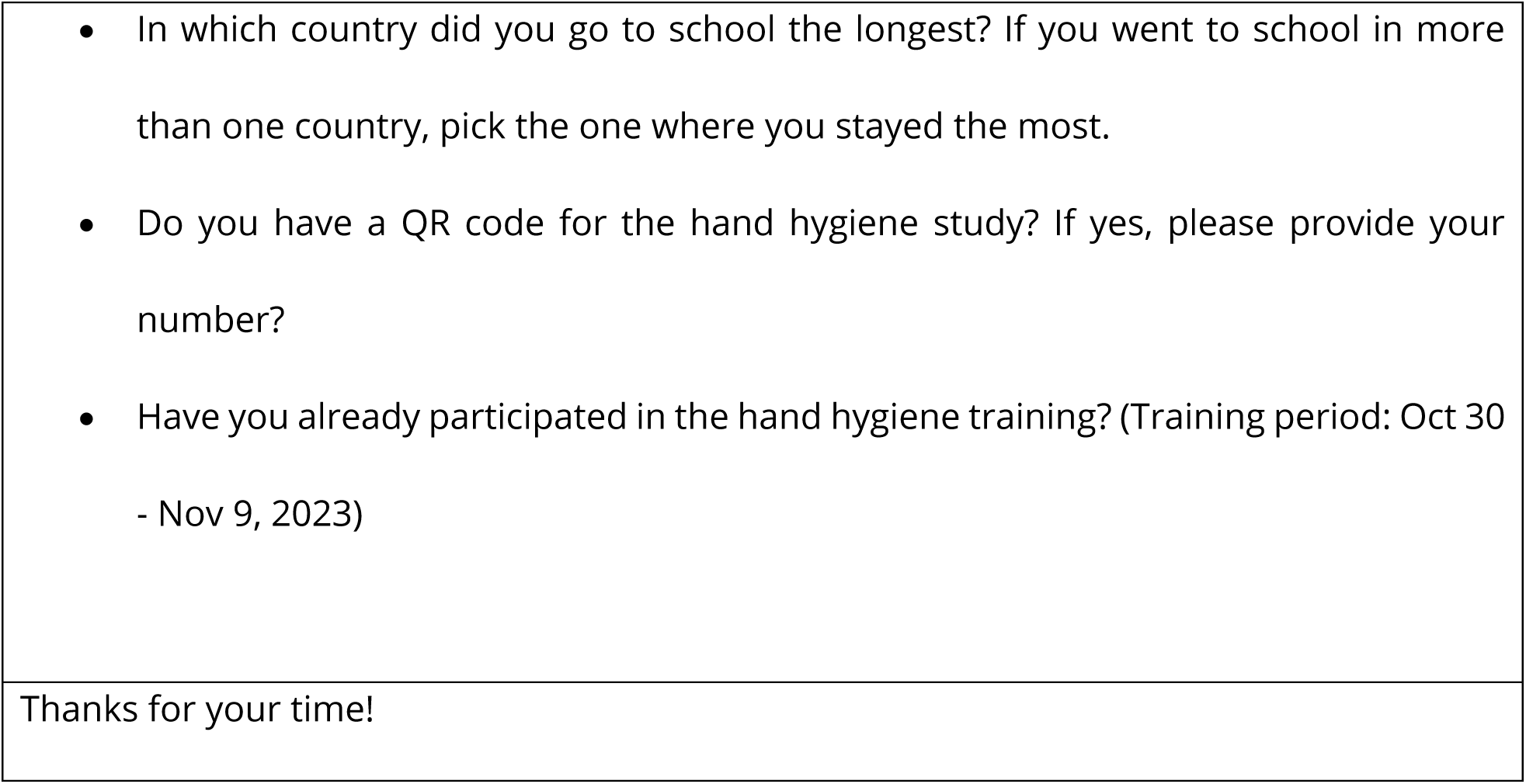
Hand Hygiene Questionnaire.

